# Advancing genetic pathogenicity prediction with three-dimensional proteoform-phenotype analysis

**DOI:** 10.64898/2026.10.07.26364649

**Authors:** Young Joo Sun, Gabriel Velez, Angela Li, Aarushi Kumar, Jennifer T. Vu, Claudia Phillips, Ditte K. Rasmussen, Jin Kyun Oh, Mauricio G. S. Costa, Stephen H. Tsang, Yang Sun, Alexander G. Bassuk, Vinit B. Mahajan

**Affiliations:** Molecular Surgery Laboratory, Byers Eye Institute, Department of Ophthalmology, Stanford University, Palo Alto, CA, USA; Department of Biomedicine, Aarhus University, Denmark; Departments of Ophthalmology, Pathology & Cell Biology, Columbia Stem Cell Initiative, Vagelos College of Physicians & Surgeons, New York, NY, USA; Jonas Children’s Vision Care, Edward S. Harkness Eye Institute, New York Presbyterian Hospital, New York, NY, USA; Programa de Computação Científica, Vice-Presidência de Educação Informação e Comunicação, Fundação Oswaldo Cruz, Riode Janeiro, Brasil; Departments of Pediatrics and Neurology and The Iowa Neuroscience Institute (INI), University of Iowa, Iowa City, IA, USA

**Keywords:** PRPH2, peripherin-2, AlphaFold, inherited renal disease, genotype-phenotype

## Abstract

Ascertaining the pathogenicity and clinical relevance of genetic mutations is a longstanding challenge across human disease. This is particularly relevant for genes with many variants of unknown significance (VUS), such as *PRPH2*-retinopathy, where complex variable phenotype patterns and limited genotype correlations hinder application of precision-medicine therapies. Recent AI-based pathogenicity tools, while powerful, may be limited by insufficient mechanistic or phenotypic training data. We established a three-dimensional proteoform-phenotype analysis framework by combining AI-generated protein structural models with clinical genomics data, in which expert consensus adjudicates model outputs through a human-in-the-loop AI approach, to evaluate 46 curated pathogenic missense variants against a background VUS pool. The framework was prospectively validated using an independently evaluated clinical patient cohort. This analysis resolved spatial biophysical signatures corresponding to distinct clinical phenotypes (i.e., retinitis pigmentosa, macular dystrophy, and pattern dystrophy) as well as complex blended presentations. Leveraging these confirmed structural signatures, we prioritized high-risk VUS. This mechanism-resolved framework can be extended to other therapeutically actionable genes to identify novel causal variants and improve diagnosis, prognosis, and better select rare, eligible patients for emerging molecular gene therapy trials.

**Plain Language Summary:** Distinguishing disease-causing mutations from benign ones remains a major barrier to precision medicine. Here, the authors combine AI-generated protein structures with clinical data and expert human adjudication to show that where a PRPH2 variant sits in three-dimensional space predicts the specific retinal disease it causes, enabling variant prioritization for gene-therapy trials

## Introduction

Next-generation sequencing (NGS) is a powerful tool for determining the molecular cause of human disease, yet its widespread use has uncovered an enormous number of alleles with unknown clinical pathogenicity.^1^ These variants of unknown significance (VUS) account for up to 40% of all identified sequence variants.^2^ Discerning benign genetic noise from a disease-causing mutation is essential for precision-health approaches, especially in rare Mendelian conditions. Establishing variant pathogenicity currently relies on family pedigrees, statistical genetics linking clinical cohorts, and *in silico* tools that analyze linear amino-acid sequences for evolutionary conservation and physicochemical properties.^3,4^ These methods, however, can fall short. A single patient may carry variants across multiple genes, multi-generational pedigree data are often unavailable, and even when a variant is deemed pathogenic, its precise clinical trajectory remains obscured.^5^ This highlights an important distinction in translational genomics: predicting whether a mutation is pathogenic is fundamentally separate from predicting which specific clinical disease it will manifest.

The clinical stakes of this distinction are nowhere more apparent than in gene therapy. Across a global landscape of more than 3,900 gene therapy trials, an expanding proportion specifically targets rare, orphan diseases.^6^ Because these patient populations are inherently small, trial design demands a delicate balance to both maximize recruitment for statistical power while ensuring diagnostic precision to avoid enrolling individuals carrying misclassified variants. Here, prediction accuracy that may be acceptable for genome-wide triage is insufficient. A tool with eighty percent accuracy is valuable for hypothesis generation across thousands of genes, but in a small trial each misassigned patient, enrolled either with a mechanistically mismatched variant or wrongly excluded, carries a cost that population-scale accuracy cannot absorb. Optimizing candidate selection without introducing confounding genotypes is a prerequisite for trial success. Because proteins, not genes, are the direct functional executors of cellular physiology, similar genomic alterations can yield strikingly different clinical phenotypes depending on the local protein structural microenvironment. Traditional low-throughput mechanistic studies have resolved this for individual variants, but physical assays are bottlenecked by cost, time, inadequate in vitro readouts, and a lack of patient-specific in vivo models, rendering them impractical for systematic personalized variant interpretation.^7,8^

Artificial intelligence now offers unprecedented opportunities to analyze human genetic variation at scale. Sequence-based missense predictors such as AlphaMissense leverage deep learning and evolutionary constraints to score variant pathogenicity, yet they operate on linear sequences and remain blind to structural mechanisms and detailed clinical phenotyping.^9–12^ Separately, structural prediction pipelines including AlphaFold and RoseTTAFold provide high-confidence 3D protein coordinates previously inaccessible to legacy tools.^9–11^ For un-crystallized proteins, these coordinates accurately describe wild-type structures but do not, in themselves, indicate whether a given variant is pathogenic or which clinical phenotype it will produce. Bridging this gap requires recognizing that computational automation and clinical curation need not be mutually exclusive. Structural and sequence-based predictors excel precisely where human curation cannot operate by processing thousands of variants with consistent, tireless pattern detection across enormous coordinate space. Human adjudicators, in turn, resolve the clinical ambiguity, phenotypic nuance, and shifting diagnostic context that a model trained without contemporaneous clinical experience cannot supply. Treating these as competing approaches, where one must substitute for the other, discards the capability each supplies where the other falls short. By synergizing the scale and consistency of AI structural modeling with expert clinical adjudication, human judgment can catch algorithmic blind spots and ground high-stakes computational predictions in real-world disease context, achieving a level of diagnostic precision that neither automated tools nor human oversight could deliver alone.^13^

Among the more than 250 genes linked to inherited retinal dystrophies, *PRPH2* is a premier archetype of extreme phenotypic heterogeneity, making it an ideal test case.^14,15^ *PRPH2* encodes peripherin-2, a photoreceptor-specific tetraspanin essential for morphogenesis and maintenance of rod and cone outer-segment discs.^7^ Its clinical presentations are so diverse that patients are frequently misdiagnosed with age-related macular degeneration, Stargardt disease, or retinitis pigmentosa (RP). Most strikingly, the *same* mutant allele can produce vastly divergent phenotypes—from localized macular pattern dystrophy to global peripheral RP—even among members of a single family.^16,17^ This variability could be shaped by trans-acting modifiers like the obligate partner ROM1, polygenic background, and stoichiometric effects.^18–20^ Four clinically divergent phenotypes recur: central areolar choroidal dystrophy (CACD), pattern dystrophies (PD), RP, and cone/cone-rod dystrophy (CD/CRD). Decoding this landscape is essential to enable rationale-based patient stratification for the emerging molecular therapies for *PRPH2* patients.

In clinical genetics, labeling a variant “pathogenic” is supported by concrete, empirical evidence: a patient presents with clear retinal degeneration captured via multimodal imaging (OCT/FAF), often backed by family pedigree segregation. Conversely, population databases like gnomAD are unphenotyped and contain asymptomatic individuals, individuals who have not yet reached the age of onset for late-stage maculopathies, or individuals with undocumented mild sub-clinical phenotypes. To address this, we developed a 3D proteoform-phenotype analysis built on a human-in-the-loop AI (HITL-AI) framework. We integrated highly characterized *PRPH2* variants with multimodal imaging phenotypes from independent patient cohorts and mapped them onto an AI-generated protein structure scaffold. Rather than introducing a black-box classifier, our approach uses high-confidence atomic coordinates, adjudicated by expert consensus, to decode phenotypic specificity across a calibration phase and an independent validation phase designed to eliminate data leakage. We show that this approach uncovers the structural rules governing disease expression far more effectively than automated pathogenicity scoring alone. We deliberately trade genome-scale breadth for mechanistic depth, and this methodology is generalizable to other complex, multi-phenotypic loci where similar translational imperatives exist and where recreating every VUS in an animal model is impractical.

## Results

### High-confidence 3D mapping overcomes the inherent limitations of sequence-based pathogenicity predictors

To systematically resolve the structural determinants of *PRPH2*-retinopathy, we first constructed a high-resolution 3D reference structural map of the human PRPH2 complex by integrating AI-generated coordinates with empirical cryo-EM structures. We then established a multi-step filtering workflow to map clinically adjudicated variants onto this spatial framework. To populate the reference map with unambiguous disease alleles, positive selection required missense variants to have definitive clinical-expert curation supported by multimodal retinal imaging or pedigree segregation. This yielded a primary discovery cohort of 46 curated pathogenic missense variants representing the full clinical spectrum (i.e., retinitis pigmentosa [RP], macular dystrophy [MD], pattern dystrophy [PD], and blended phenotypes) which was complemented by an independent, prospectively evaluated clinical validation cohort (n = 31). To maintain analytical rigor within this 3D framework, two distinct variant classes were systematically excluded. First, non-missense mutations, including frameshift, nonsense, and splice-site variants, were removed because large structural truncations or null alleles disrupt subunit stoichiometry and global protein expression, rendering single-residue 3D biophysical metrics (e.g., ΔΔG, atomic depth, and solvent accessibility) structurally uninterpretable. Second, uncurated variants from general population repositories like gnomAD were excluded from serving as a “benign control” baseline. Because population databases contain unphenotyped individuals, non-penetrant carriers, and individuals who have not yet reached the age of onset for late-stage maculopathies, treating uncurated population entries as true controls introduces false-negative noise into singular-class biophysical profiling.

A ClinVar database search initially identified 503 *PRPH2* variants. Rather than relying on conventional binary classifications that require a comparative “benign” control cohort, we adopted a singular-class, intrinsic biophysical profiling framework. Because treating uncurated gnomAD variants as a true benign baseline introduces severe confounding noise and false-negative controls, population-level frequencies were excluded from our analytical framework. The computational workflow was trained exclusively on the physical and structural footprints of clinically curated disease alleles. Instead of measuring how rare a variant is in the general population, this framework determined disease risk by examining whether a mutation directly disrupts PRPH2 structural integrity.

To construct a high-confidence, unambiguous 3D spatial reference map, we systematically excluded variants with highly conflicting clinical records, those reported to simultaneously cause multiple divergent phenotypes without consensus, and those lacking peer-reviewed clinical data. This stringent filtering pipeline yielded 164 highly characterized coding variants with clear, consensus clinical phenotypes to serve as our baseline dataset for the primary 3D proteoform-phenotype analysis (**Extended Data Tables 1–4**).^21^ Of these, 93 were pathogenic and 71 were VUS. No clear genotype-phenotype correlation was observed based on 1-dimensional sequence alignment alone (Figure **1a–b** and Supplemental Figure **3**). Within the 93 pathogenic variants, 46 were missense changes: three were linked to CACD (6.5%); ten to PD (21.7%); 22 to RP (47.8%); five to CD/CRD (10.9%); and six to multiple phenotypes (13.0%; Figure **1b–c** and **Extended Data Table 2**).^5,22–75^ We analyzed these 46 pathogenic missense *PRPH2* variants using AlphaMissense.^12^ The machine learning model misclassified several entries, predicting eight of these known pathogenic variants to be either benign (6 variants) or ambiguous (2 variants). Ensemble predictors like MetaRNN and BayesDel similarly exhibited high sensitivity for generalized pathogenicity, yet they still suffered from notable false-negative rates on *bona fide* pathogenic variants (Figure **1c**).^76,77^ This underscores that predicting the pathogenicity of *PRPH2* missense variants can remain challenging for generalized sequence-based predictors (**Extended Data Table 5**). Because automated sequence predictors were trained primarily on population variant frequencies, AlphaFold-based structural chemistry, and broad ClinVar pathogenicity annotations, they are inherently blind to inheritance patterns and specific clinical phenotypes.^12^ To address these limitations, human structural and phenotypic adjudication was integrated, an emerging approach for clinical AI research.

**Figure 1.**
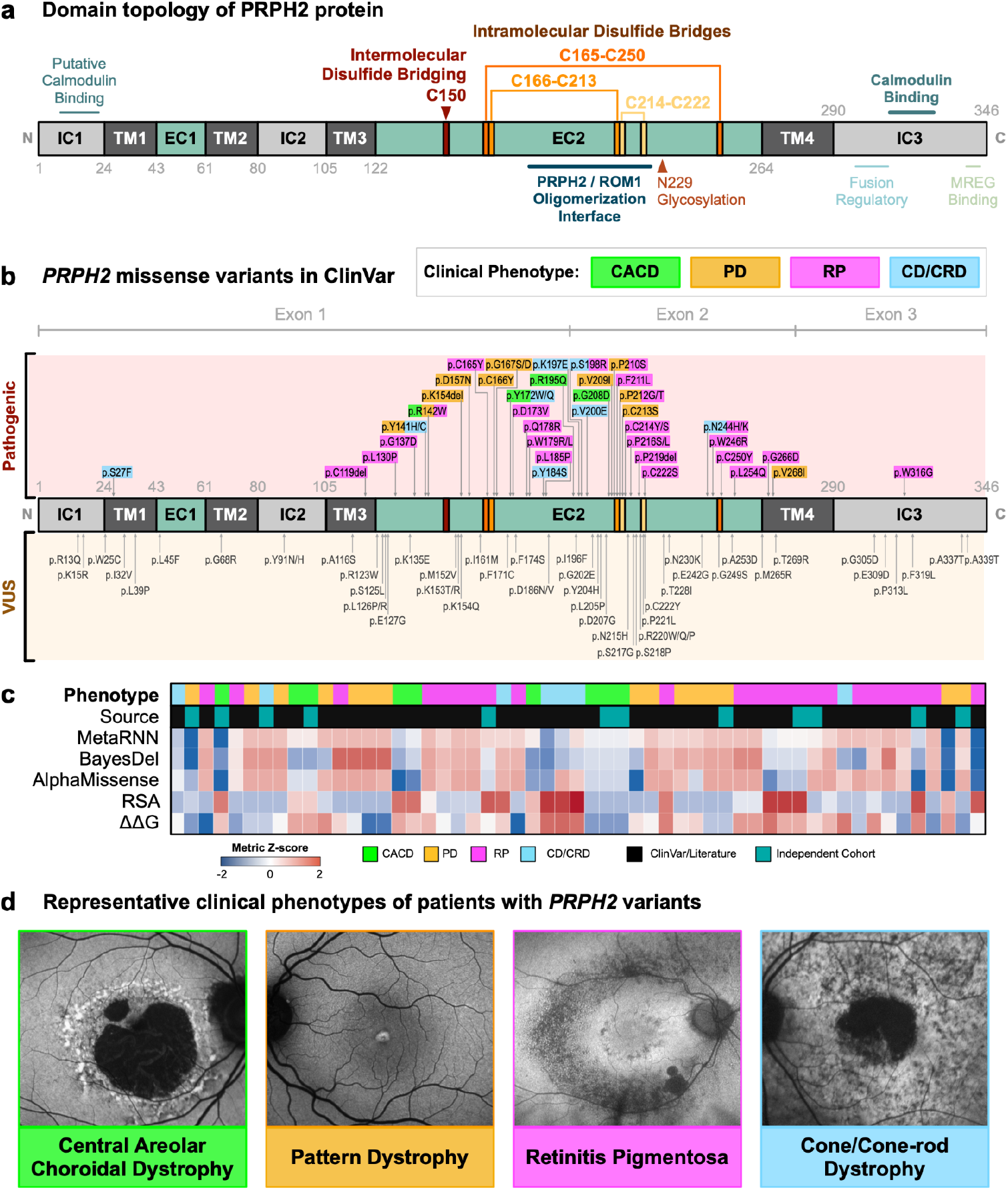
PRPH2 disease variants in 1-dimension do not correlate with clinical phenotypes: (a) Linear schematic (primary structure map; domain topology map) showing the structural and functional domains of the PRPH2 protein. The seven key cysteine residues involved in intermolecular interaction (Cys150; brown) and in intramolecular folding (Cys165, Cys166, Cys213, Cys214, Cys222, Cys250; orange pallets) are shown. The cysteine residues forming intramolecular disulfide bridges are paired. The N-glycosylation site, Asn229, is shown in light brown. Key functional regions are indicated: calmodulin binding, oligomerization interface with ROM-1 and/or PRPH2, fusion regulation, and melanoregulin (MREG) binding. (b) Previously reported PRPH2 missense variants are mapped onto the domain topology map. Pathogenic variants and VUS are shown above and under the map, respectively. Pathogenic variants shown in green, yellow, magenta, and cyan boxes were identified in the patients displaying CACD, PD, RP, and CD/CRD, respectively. (c) Multiparametric heatmap tracking 96 annotated single-amino-acid substitutions across clinical, structural, and computational prediction axes. Variants are stratified by their clinical phenotype (CACD, green; PD, orange; RP, magenta; CD/CRD, cyan) and cohort discovery source (curated ClinVar/literature variants, black; independent cohort, teal). Continuous metric rows, comprising MetaRNN, BayesDel, and AlphaMissense pathogenicity scores alongside relative solvent accessibility (RSA) and SDM thermodynamic folding stability (ΔΔG), are standard-normalized and color-ramped by Z-score metric intensity. Blue denotes a negative Z-score deviation, while red denotes a positive Z-score shift (over-representation/increased solvent exposure/severe destabilization). (d) Representative clinical fundus autofluorescence images acquired from retinal dystrophy patients with PRPH2 variants harboring CACD, PD, RP, and CD/CRD, respectively.

We generated a 3D proteoform-phenotype map by combining the genetic and phenotypic parameters of the 46 pathogenic missense variants with structure-function data from human PRPH2, projecting them onto a 3D structural model obtained from the AlphaFold database (Supplemental Figure **1a–b**).^9^ We utilized the AlphaFold model to test the applicability of our framework to inherited disease-causing genes lacking experimentally resolved structures, benchmarking it against AlphaMissense. During the course of our study, the cryogenic electron microscopy (cryo-EM) structure of wild-type human PRPH2 in complex with ROM1 (PDB ID: 7ZW1) was solved, so we utilized this structure to assess the accuracy of the AlphaFold model.^78^ AlphaFold accurately recapitulated structural features detected in the cryo-EM structure, including the seven conserved cysteines in the EC2 domain that form three distinctive intramolecular disulfide loops (Supplemental Figure **1c**), which were poorly resolved by legacy non-AI homology modeling tools. Notably, AlphaFold provided a complete structural model of human PRPH2—encompassing the four transmembrane (TM) domains, two extracellular domains (EC; intradiscal in rods), three intracellular domains (IC; cytoplasmic in rods). While the published cryo-EM constructs physically span residues 2–346, local conformational flexibility in the distal intracellular C-terminus results in a lack of interpretable electron density, causing empirical atomic coordinates to truncate prematurely around residues 320–325. AlphaFold effectively overcame this experimental limitation by resolving a fully coordinated, continuous backbone model through to the terminal residue (Gly346). This permitted the mapping and analysis of pathogenic patient variants residing in this functionally vital, flexible C-terminal tail that were otherwise untraceable in the experimental structural coordinates (Figure **1a** and Supplemental Figure **2**).^79^

### Spatial core sequestration, rather than baseline thermodynamic instability, dictates PRPH2 pathogenicity

To isolate the general properties driving PRPH2 pathogenicity, variations in protein biophysical attributes, including calculated thermodynamic stability changes (ΔΔG), relative solvent accessibility (RSA), and native atomic depth, were evaluated. Interestingly, global calculated thermodynamic shifts failed to differentiate established pathogenic drivers from the background VUS pool, with both groups displaying nearly identical mean destabilization profiles (pathogenic: −0.711 ± 1.26 kcal/mol; VUS: −0.778 ± 1.28 kcal/mol; p = 0.633; Figure **2a**). In contrast, 3D topological metrics revealed robust, statistically divergent positioning trends between the two tracks. Confirmed pathogenic mutations were highly sequestered within the interior structural volume of the PRPH2 matrix, yielding a significantly lower mean relative solvent accessibility (pathogenic: 23.7% vs. VUS: 37.0%; p = 0.012) and a significantly deeper native atomic anchoring depth (pathogenic: 5.62 Å vs. VUS: 4.71 Å; p = 0.018; Figure **2a**).

**Figure 2.**
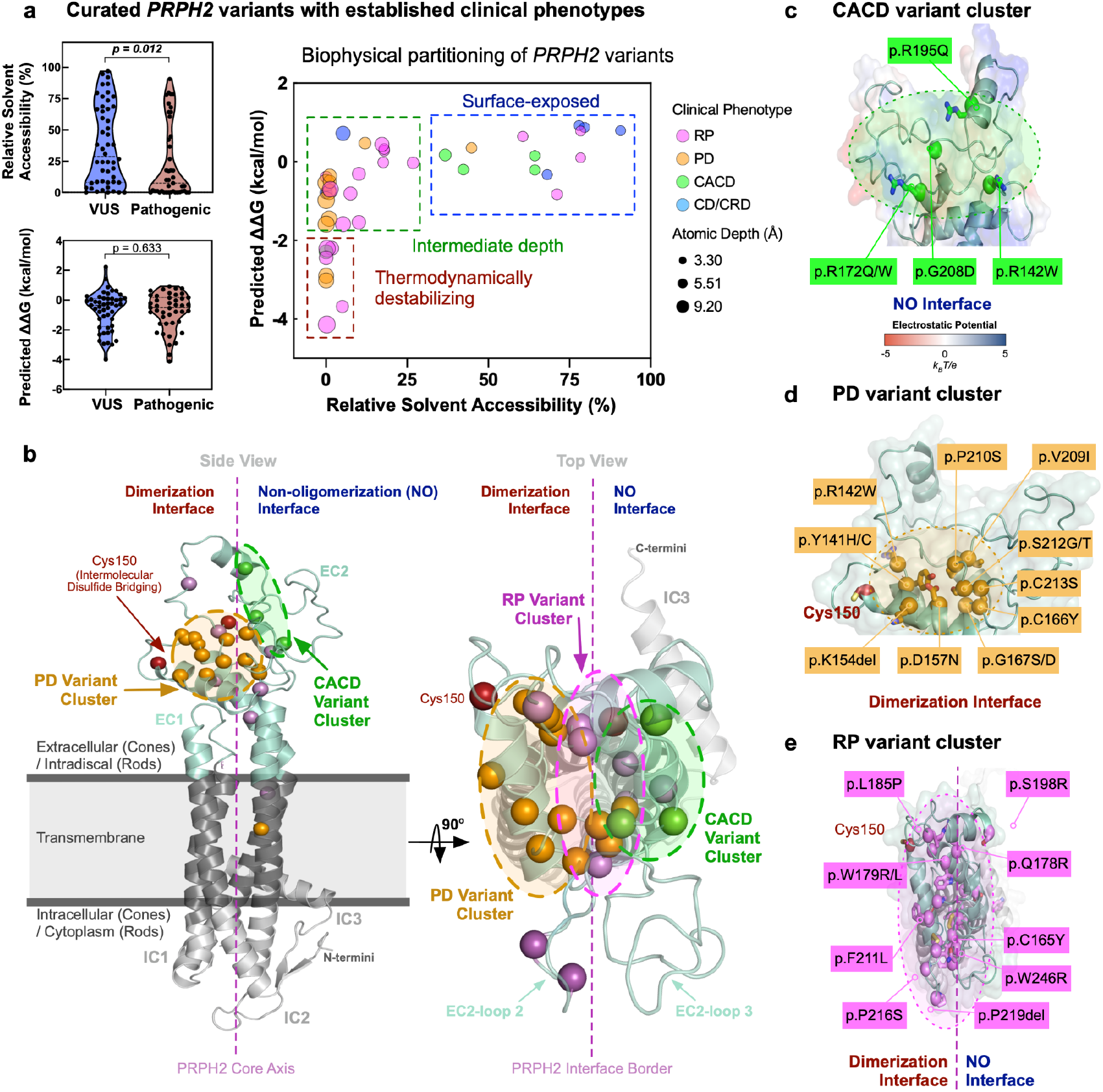
Mapping pathogenic missense variants in a 3-dimensional AI generated protein structure reveals clustering by clinical phenotype: (a) Multiparametric partition profiling of PRPH2 variants. Independent violin plots (left) illustrate the continuous distribution of relative solvent accessibility (RSA) and SDM-predicted thermodynamic folding stability shifts (ΔΔG, kcal/mol) compared between VUS and established pathogenic mutations (p-values derived via two-tailed Mann-Whitney U testing). Central scatter visualization maps the synchronous covariance of native structural parameters across the 96 annotated variants. Dashed geometric gates delineate three distinct physical phenotype zones: a thermodynamically destabilized interior core region (RSA < 30%, ΔΔG < −2.0 kcal/mol), an intermediate boundary depth layer (RSA 30-40%), and a neutral surface-exposed domain (RSA > 40%, ΔΔG ~ 0 kcal/mol). Data points are scaled by calculated atomic depth (Å) and color-coded by associated clinical phenotype. (b) Ribbon-trace diagram of the PRPH2 AlphaFold model. Side view (left) highlighting the position of known pathogenic missense mutations in PRPH2 (denoted as spheres). Mutations are colored according to their respective clinical phenotype (CACD, green; PD, orange; RP, magenta) and dotted spheres denote the respective mutation clusters. The dimerization and non-oligomerization interfaces (left and right, respectively) are delineated by the vertical red dotted line. Top view (right) highlights the position of the RP mutation cluster (magenta) as well as variants clustering in EC2-loop 2. (c) Surface diagram highlighting CACD-associated pathogenic missense variants in PRPH2 on the non-oligomerization interface (i.e., CACD cluster). The transparent surface is colored continuously by local electrostatic potential calculated via the Adaptive Poisson-Boltzmann Solver (APBS) across a scale of −5.0 kBT/e (electropositive, red) to +5.0 kBT/e (electronegative, blue). White regions denote neutral surface patches. (d) Surface diagram highlighting PD-associated pathogenic missense variants in PRPH2 on the dimerization interface (i.e., PD cluster). Proximity to the intermolecular disulfide bridging C150 (red sphere) is demonstrated. (e) Surface diagram highlighting RP-associated pathogenic missense variants in PRPH2 (magenta spheres).

Analysis of curated variants based on their primary anatomical presentation, contrasting peripheral rod-dominant dystrophies against a combined spectrum of macular-predominant diseases affecting the central retina, revealed a sharp biophysical segregation (Figure **2a**). Severe thermodynamic destabilization (ΔΔG < −2.0 kcal/mol) was localized to deeply buried architectural residues (RSA < 15%) and was highly predictive of an RP phenotype. Conversely, surface-exposed variants (RSA > 25%) demonstrated near-neutral thermodynamic profiles (ΔΔG ~ 0 kcal/mol) yet strongly associated with the aggregated macular-predominant spectrum (PD, CACD, CD/CRD). This dichotomy suggests that central retina-affecting phenotypes are driven primarily by disruptions to outer-surface intermolecular assembly rather than monomeric unfolding. Together, these results demonstrate that the spatial localization of an alteration relative to the protein core matrix, rather than its unconstrained, baseline calculated thermodynamic penalty, may serve as the primary structural signature of disease-driving variation in PRPH2. Leveraging these statistically confirmed topological constraints (i.e., RSA < 15% and Depth > 5.5 Å), we successfully prioritized a high-risk subset of 11 core-nested VUS entries (including p.G68R, p.L126P, and p.W174S; **Extended Data Table 3**) that biophysically mimic known pathogenic trajectories, suggesting their strong potential for clinical reclassification.

To evaluate whether broader clinical phenotypes share common spatial mechanisms, we aggregated the 46 pathogenic missense variants based on their primary anatomical presentation. This macro-level categorization revealed a striking, spatially distinct structural segregation across the 3D proteoform (Figure **2b**). Variants driving peripheral retinal degeneration (RP) preferentially occupied the densely packed, internal structural core of PRPH2, a configuration strongly correlated with compromised folding stability. In contrast, central retina-affecting variants (PD, CACD, CD/CRD) were significantly enriched on the solvent-exposed outer surface boundaries and functional extracellular domains of the protein. This distinct outer-surface clustering suggests that macular-predominant phenotypes are driven primarily by disruptions to intermolecular and higher-order macromolecular interactions rather than the destabilization of individual monomeric units. CACD-associated missense variants (e.g., p.R172Q, p.R172W, p.R195Q, and p.G208D) clustered on an extracellular lobe that does not participate in the standard oligomerization of PRPH2, identifying a clear non-oligomerization (NO) interface (Figure **2c**).^80^ The NO interface faces the intradiscal space in rod photoreceptors, whereas in cone photoreceptors, this interface is exposed to the extracellular environment and accessible to separate molecular partners. This structural difference may explain the cone-dominated, central vision phenotypes unique to CACD. On the opposite side of the CACD cluster, every missense variant harboring the PD phenotype clustered on the dimerization interface, which is essential for both PRPH2 homodimerization and PRPH2/ROM1 heterodimerization (Figure **2d**).^78,80^ These PD variants affect the cysteine residues that establish vital intramolecular disulfide bridges or alter amino acids in close spatial proximity to them. These residues are crucial for dimerization and subsequent higher-order oligomerization of PRPH2 required for proper outer segment disc morphogenesis.^72,78,80,81^

RP-associated missense mutations appeared dispersed throughout PRPH2 with no obvious pattern when viewed 1-dimensionally (Figure **1b**). Yet, through a 3D lens, these variants clustered in two structural regions: the EC2-loop 2 and the EC2 core region separating the NO-and dimerization-interfaces (Figure **2e**).^78,80^ EC2-loop variants (p.C214S, p.C214Y, p.P216S, p.P216L, and p.C222S) mapped to a region formed by a disulfide bridge between C214 and C222 that participates directly in PRPH2 dimerization. Structural modeling of these EC2-loop RP mutants indicates that structural substitutions of these cysteines and prolines disrupt rigid local secondary structures, altering the conformation and stability of the loop. The remaining 15 RP mutants mapped directly to the EC2 core regions, which are critical for PRPH2 folding and stability (Figure **2e** and **Extended Data Table 6**).^82^ Our findings support prior *in vitro* studies showing that p.L185P prevents oligomer formation, leading to protein instability and degradation, indicating these destabilized PRPH2 variants likely result in a haploinsufficient RP phenotype. Overall, central retina-affecting variants (e.g., CACD and PD) localize to peripheral, surface-exposed surfaces where they impair intermolecular interactions, while peripheral retina-affecting variants (i.e., RP) are anchored within the structural core, destroying monomeric folding integrity.^83,84^ Taken together, this PRPH2 proteoform-phenotype analysis reveals that different clinical phenotypes cluster distinctively in 3D space, implying separate structural pathologies.

### Topological modeling resolves ambiguous VUS classifications and explains blended phenotypic presentations

Next, we verified the predictive potential of the PRPH2 3D structure-phenotype framework within an independent clinical cohort. Screening 4,945 retinal dystrophy (RD) patients from two tertiary academic eye centers identified 31 individuals (0.63%) harboring a total of 23 *PRPH2* variants (48% female; age range 12–79 years; Supplemental Table 1). The identified *PRPH2* missense variants included six pathogenic entries, two novel entries, and five VUS. Novelty was defined as a complete absence of the allele from all major reference repositories, including ClinVar, the gnomAD population database, and the indexed biomedical literature via PubMed at the time of our cohort assembly. Non-missense changes included four nonsense mutations (three pathogenic, one VUS), five frameshifts (two pathogenic, three novel), and one novel insertion (Supplemental Table 1). Clinical phenotypes comprised CACD (n = 12), PD (n = 9), RP (n = 4), and CD/CRD (n = 6; Supplemental Figure **4-7**). The majority of patients were heterozygous for a single *PRPH2* change, with the exception of two individuals who each co-harbored a pathogenic variant and a background VUS (Supplemental Table 1). Most patients (74%) harbored no variants for other retinal disease genes (**Extended Data: Case Series**).

To assess how well the 3D structure-phenotype framework predicts clinical presentation, we mapped the six pathogenic missense variants identified within our patient cohort (Figure **2**). Notably, one of the pathogenic variants (p.R172W) in our cohort was inaccurately flagged as ‘benign’ by AlphaMissense, yet the clinical phenotype of every pathogenic-variant patient in our repository matched the predictions of our structural framework.^12^ Patients harboring the known pathogenic variants p.R142W, p.R172W, and p.G208D (Supplemental Figure **4a–c**) presented with clinical features of CACD, a macular dystrophy localized to the central retina, retinal pigment epithelium (RPE), and choriocapillaris.^5,28,85,86^ These variant residues cluster tightly around a patch of basic residues on the NO interface (the CACD cluster; Figure **2c**), corroborating our proteoform-phenotype map. Patient P9 harbored p.C213S, a pathogenic variant tied to PD, a disease spectrum defined by reticular macular pigment deposition.^24^ On our 3D framework, this variant maps directly to the pathogenic PD cluster, and clinical examination of P9 revealed classic fleck-like lesions matching STG-PD (Supplemental Figure **5a**). Another pathogenic variant, p.Y141C, also maps to the PD cluster, yet it displays a highly variable RP phenotype, even within families.^36–38,69,87–91^ Mutation of the same residue to a histidine (p.Y141H) leads to PD-spectrum phenotypes like adult vitelliform macular dystrophy (AVMD). This phenotypic divergence is explained by the proximity of Tyr141 to Cys150, a region controlling oligomerization kinetics. In the case of p.Y141C, introducing an ectopic cysteine prompts abnormally large PRPH2 oligomer formation in mouse models, whereas p.Y141H impairs assembly, mimicking other PD-associated variants.^18,36^ In our cohort, patient P10 co-harbored *PRPH2* p.Y141C and the pathogenic *ABCA4* variant p.R2030Q, presenting with a cone-rod dystrophy involving the posterior pole; here, the companion *ABCA4* mutation likely acts as an epistatic phenotype modifier (Supplemental Figure **7a**). The clinical presentations of all other pathogenic variants in our cohort were accurately predicted (Supplemental Figure **5**). Taken together, these results demonstrate that the 3D structure-phenotype framework successfully links genotypes to specific clinical phenotypes, solidifying the hypothesis that variants impacting the same localized structural interface yield congruent clinical presentations.

While automated sequence predictors like AlphaMissense offer continuous scores for generalized pathogenicity, they lack the structural and phenotypic context required to predict distinct clinical disease manifestations. To test this, we applied the 3D structure-phenotype framework to patients in our cohort with a *PRPH2* missense VUS (Supplemental Table 1). Of the 5 VUS in our patient cohort, three were predicted pathogenic, one was ambiguous, and the other was benign based on AlphaMissense.^12^ Some variants were independently reported in retinal dystrophy cohorts (e.g., P12: p.K135E; P13: p.S218P; and P14 and P15: a mother/daughter pair with p.G305D; Figure **3a**).^24,69,92^ Most notably, the 3D structural analysis successfully resolved the false-benign classification of the VUS p.K135E. Because automated sequence-based algorithms rely heavily on linear evolutionary alignment, AlphaMissense erroneously classified p.K135E as ‘benign’ (score: 0.1714). In contrast, our 3D framework mapped residue Lys135 directly into a highly constrained structural loop cluster mandatory for stable secondary structural interactions, accurately aligning the severe retinal dystrophy confirmed in our patient (Figure **3**). As predicted, clinical features of P12 included a well-demarcated area of central macular atrophy with prominent paracentral hyperfluorescent rings characteristic of CACD (Figure 3c). p.S218P of P13 affects the EC2-loop 2, near the RP-associated cluster (p.P216S and p.P216L) and likely affects the loop’s rigidity (Figure **3d**). Clinical assessment of P13 showed widespread retinal atrophy with superior lacunar degeneration and peripheral pigmentary changes (Figure **3e**); full-field electroretinography confirmed late-stage RP. Finally, the p.G305D VUS mapped to the IC3 domain near the calmodulin binding motif regulating membrane fusion (Figure **3f**), where non-missense pathogenic variants in this region that disrupt structure-function (e.g., p.S301fs, p.S301Rfs*22, p.E302*, p.W316fs, and p.W316*) are associated with PD, specifically the AFVD type (**Extended Data Table 4**).^93,94^ Likewise, the p.G305D variant is expected to affect membrane fusion because it reduces loop flexibility and introduces a negative charge to IC3. Clinically, P14 and P15 displayed hyperfluorescent vitelliform-like lesions consistent with AFVD (Figure **3g**). Interestingly, there were three VUS found in this same region in patients with PD (p.V312fs, p.E321*, and p.K235Rfs*35; **Extended Data Table 4**).

**Figure 3.**
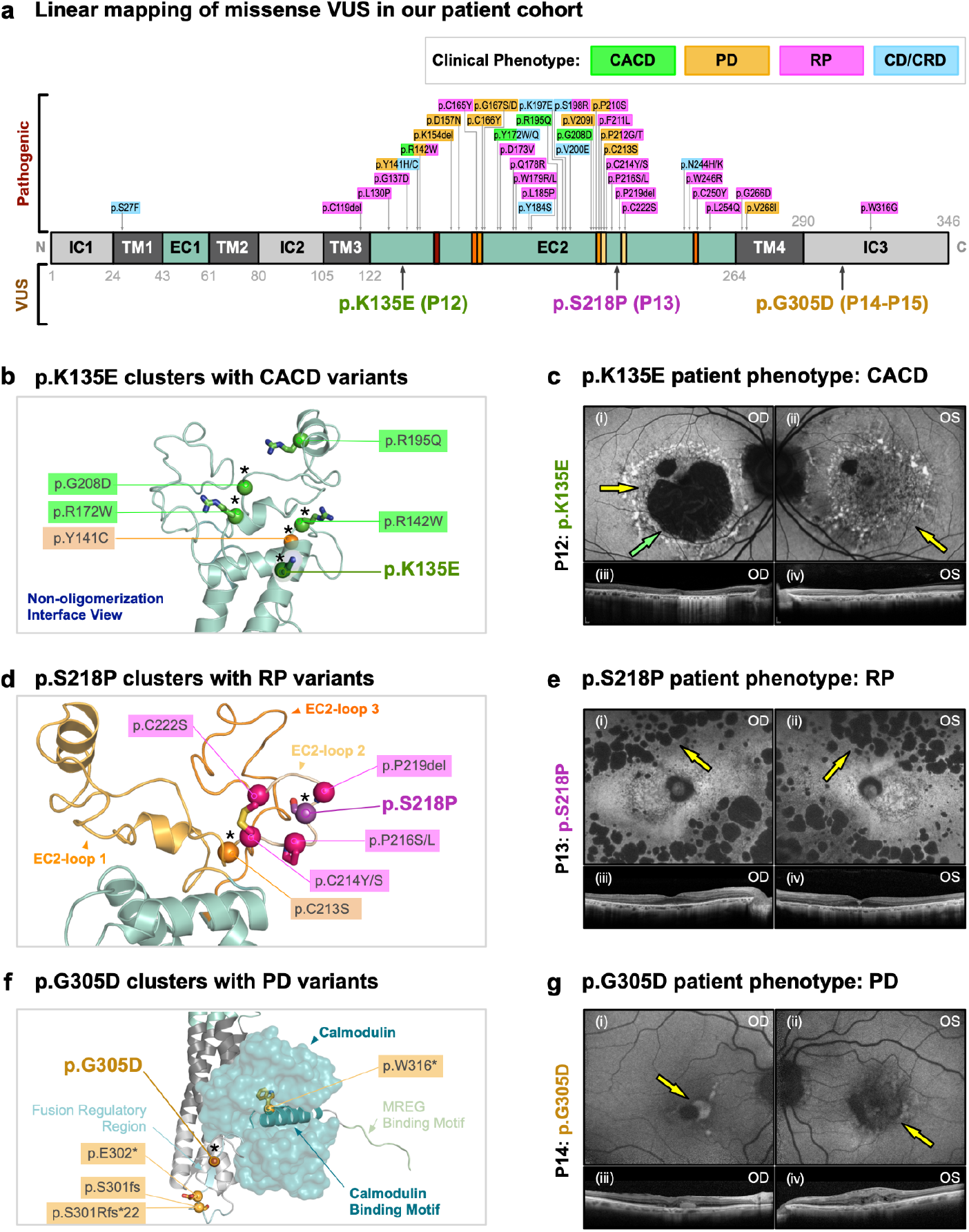
3D structure-phenotype framework predicts clinical phenotypes of missense VUS by their tertiary-structural location: (a) PRPH2 missense VUS that were found in our patient cohort are mapped on the primary structure (linear schematic) of the PRPH2 protein. Pathogenic variants and our patient cohort’s VUS are shown above and under the map, respectively. Pathogenic variants shown in magenta, orange, green, and cyan boxes were identified in the patients displaying RP, PD, CACD, and CD/CRD, respectively. Variants identified in our patient cohort are underlined. (b) p.K135E clustered at the non-covalent oligomerization interface with other PRPH2 pathogenic missense variants (green box) identified in CACD patients. (c) Patient 12 (P12) with the p.K135E variant displayed CACD phenotypes, including speckled pattern of fundus autofluorescence (yellow arrows) and a central area of no autofluorescence corresponding to severe RPE atrophy (green arrow). (d) p.S218P clustered at the EC2-loop 2 with other PRPH2 pathogenic missense variants (magenta box) identified in RP patients. (e) Patient 13 (P13) with p.S218P variant displayed RP phenotypes, including discrete areas of hypo-autofluorescence indicating RPE loss in the mid peripheral retina in both eyes (yellow arrows). (f) p.G305D clustered at the IC3 domain with other PRPH2 pathogenic variants (yellow box) identified in PD patients. The IC3 domain contains the fusion regulatory region, calmodulin binding motif, and MREG binding motif. Model of Calmodulin (deep green surface model; PDBID: 2HQW) bound to PRPH2’s calmodulin binding motif is shown. (g) Patient 14 and 15 (P14-15) with p.G305D variant displayed PD (AFVD) phenotypes including a small, hyper-autofluorescent foveal lesion in the right eye and central hypo-autofluorescence in the left eye (yellow arrows). P15 images not shown. Asterisks denote variants identified prospectively within our clinical cohort (including the 6 novel variants and 6 VUS).

We next expanded our study to 52 missense VUS in the ClinVar database (Figure **4**). Five were identical to VUS in our first patient cohort. Nine missense ClinVar VUS mapped to the NO-interface. Interestingly, patients with these VUS reportedly displayed both CACD features (p.R123W, p.S125L, and p.K135E) and PD features (p.S125L, p.E127G, p.K135E, p.G202E, p.Y204H, p.L205P, p.D207G, and p.E242G), which are both central retinal phenotypes (Figure **4a**, **Extended Data Table 3**). Combined with pathogenic missense variants at the NO-interface, our 3D structure-phenotype framework identified a pattern: the variants in this region were disrupting natural charged-patches at the NO-interface (Figure **4b**): loss of basic charge was associated with CACD (e.g., p.R123W, p.R172W, and p.R195Q) and disruption of acidic charge was associated with PD (e.g., p.E127G, p.D207G, and p.E242G). This suggests that the NO-interface of PRPH2 may play a crucial role through its charged-patches, specifically in cone photoreceptors, which are concentrated in the central retina, and that each charge may play preferential roles in CACD or PD pathology.

**Figure 4.**
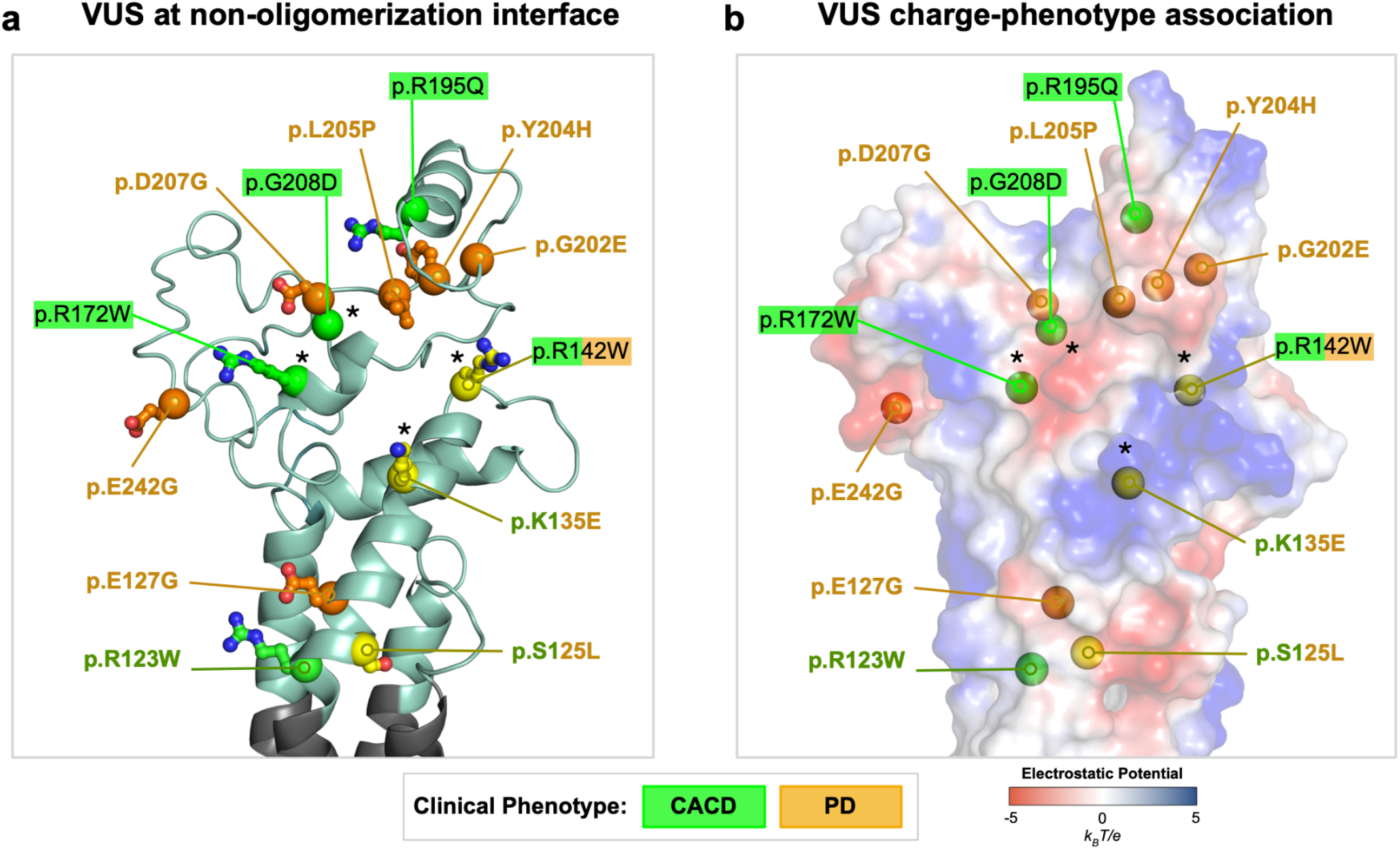
Structural analysis of pathogenic variants and VUS at NO-interface reveals surface charge distinguishes central retinal phenotypes: (a) Previously reported PRPH2 VUS and pathogenic CACD variants (sphere-stick model) mapped at the non-oligomerization interface. Variants are denoted by the following labels: VUS in PD patients, orange underline; VUS in CACD patients, green underline; VUS found in both CACD and PD patients, yellow sphere-stick models and corresponding green/orange label; pathogenic CACD variants, green box; and pathogenic CACD/PD variants, green/orange box. (b) Vacuum-electrostatics surface model of PRPH2 confirms a trend that loss of basic charge variants is associated with CACD (e.g., p.R123W, p.R172W, and p.R195Q) and disruption of acidic charge variants is associated with PD (e.g., p.E127G, p.D207G, and p.E242G). The surface is colored continuously by local electrostatic potential calculated via the Adaptive Poisson-Boltzmann Solver (APBS) across a scale of −5.0 kBT/e (electropositive, red) to +5.0 kBT/e (electronegative, blue). White regions denote neutral surface patches. Asterisks denote variants identified prospectively within our clinical cohort (including the 6 novel variants and 6 VUS).

To evaluate the versatility of the 3D proteoform analytical framework, we investigated patients within our cohort who presented with multiple *PRPH2* variants. This sub-cohort included a familial pedigree (maternal aunt P24 and nieces P25 and P26) co-harboring p.Y91N and p.I104Vfs*34, as well as an isolated case (P16) presenting with concurrent p.G208D and p.T228I missense substitutions. For patients P24, P25, and P26, pedigree tracking indicated that the variants reside in *cis*. The insertion of the truncating frameshift (p.I104Vfs*34) within transmembrane domain 3 (TM3) is predicted to prompt nonsense-mediated mRNA decay or induce severe structural collapse of the local alpha-helical bundle. This null allele effectively dominates the clinical phenotype, manifesting as a CRD regardless of the companion p.Y91N passenger variant. In contrast, patient P16 harbored two distinct missense variants (p.G208D and p.T228I). The p.G208D mutation maps directly to a high-confidence pathogenic structural cluster located on the non-covalent oligomerization interface, a region heavily linked with CACD (Supplemental Figure **8a**).^25,26^ Concurrently, the p.T228I variant alters a highly conserved residue immediately adjacent to the vital N-glycosylation consensus site at position Asn229.^2,4^ Because the loss of glycosylation at Asn229 is known to impair higher-order disk-rim oligomerization, our 3D framework rationalized that p.T228I acts as a phenotypic modifier. Consequently, the framework predicted a combined, blended clinical presentation. Multimodal clinical imaging aligned with this dual structural etiology: fundus photography revealed bilateral, diffuse yellow pisciform deposits classic of STG-PD, while FAF captured the sharply demarcated speckled macular atrophy pathognomonic of CACD (Supplemental Figure **8b**). Finally, there were 9 frameshift and nonsense variants (P19– P30; Supplemental Table 1) subject to nonsense-mediated decay or unstable, misfolded, and functionally inactive configurations without a consistent phenotype as previously reported (**Extended Data Table 4**, Supplemental Figure **3**). There was insufficient genetic and proteoform information to infer clear pathogenicity where phenotypes may have been modified by drugs or genes affecting the macula in three patients with novel missense and insertion variants (P17, P18, P31; **Extended Data: novel variants analysis**). As further patient mutations and phenotypes are identified, they can be integrated to strengthen the 3D structure-phenotype framework.

## Discussion

Recent advancements in AI-driven clinical genetics tools offer improved pathogenicity classification, yet significant diagnostic challenges remain.^12^ Inherited retinal dystrophies represent a prime example of genotypically heterogeneous diseases where traditional linear genotype-phenotype correlations may not be able to resolve complex genotype-phenotype associations, thereby slowing clinical trial recruitment and therapeutic development. In this study, a three-dimensional proteoform-phenotype analysis was deployed to elucidate these long-standing uncertainties, evaluating 164 highly characterized variants from the ClinVar database alongside six novel missense alterations identified within our institutional cohort of 4,945 retinal dystrophy patients. A cohort is defined as independent based on its patient population and clinical datasets, not solely on the exclusion of known genetic alleles.^95^ Demonstrating that a variant from the ClinVar database consistently maps to the exact same highly specific multi-modal imaging phenotype in an entirely separate patient from an independent academic medical center serves as an authentic replication of findings.^96^ This external validation step confirms the clinical penetrance and reproducibility of our structural zones without introducing the data leakage or inflated performance metrics common to models validated on internal subdivisions of a shared dataset.^97^

The human-in-the-loop step proves most valuable in cases where expert consensus diverged from the conclusion a structural perturbation score alone would suggest. In some instances, a substantial predicted structural change proved inconsistent with the observed clinical phenotype. In others, a modest predicted perturbation produced a disproportionate phenotype, attributable to its position within a critical functional region. Such cases underscore that automated structural scores and mechanistically grounded, phenotype-consistent classification are not interchangeable, a key distinction motivating this framework. Looking forward, the same HITL-AI architecture provides a natural framework for fusing additional orthogonal data streams into variant adjudication as they mature. Emerging high-throughput functional assays, multiplexed variant-effect measurements, and patient-derived cellular and animal models could supply experimental readouts that expert adjudicators integrate alongside structure and clinical phenotype where the model resolves the structure and humans integrate the broader biological system. This positions HITL-AI not merely as an interpretive tool but as a translational engine that keeps computational predictions grounded in biological reality, with relevance extending beyond any single disease gene.

Mapping these disease-causing missense variants in 3D space revealed that mutations causing CACD, PD, and RP cluster into distinct, mutually exclusive regions of the EC2 domain (Figure **2**). Integrating prior biophysical insights from a handful of experimentally validated PRPH2 mutants, this spatial architecture explains why most macular dystrophy patients harbor variants that disrupt higher-order macromolecular assembly while preserving core monomeric folding stability.^35,36,98^ Identifying these exact coordinates has prognostic value; core-nested alterations drive rapid protein destabilization, degradation, and a subsequent haploinsufficient loss of functional PRPH2 dimers that clinically manifests as severe, progressive peripheral rod degeneration (RP). Conversely, surface-exposed focal macular dystrophy variants (CACD or PD) alter localized interaction boundaries, exerting a dominant-negative effect on disc morphogenesis while leaving the underlying monomer intact.^7,35^ Our 3D structure-phenotype framework introduces two mechanistic insights to the PRPH2 landscape. First, patients presenting with the adult vitelliform macular dystrophy (AVMD) subform of PD exhibit a striking mutational enrichment within the intracellular 3 (IC3) domain, suggesting that structural modifications to this localized tail impair vital calmodulin-binding and membrane fusion kinetics (Figure **3f–g**). Second, the phenotypic bifurcation observed at the non-oligomerization (NO) interface is strictly governed by electrical charge properties; a targeted loss of basic charge drives a cone-dominated CACD presentation, whereas the disruption of adjacent acidic patches selectively precipitates PD. This observation aligns with established biophysical paradigms where localized electrostatic surface potentials function as the primary arbiters of macromolecular self-assembly and disk-rim structural packing.^99,100^

Our 3D proteoform framework provides the necessary biophysical foundation to understand how background genetic and environmental modifiers impact specific patient populations. By establishing a highly curated spatial map, we demonstrate that mutations residing on external macromolecular interfaces are uniquely sensitive to minor biochemical fluctuations in their interacting protein partners. While our primary structural mapping utilized a highly accurate AI-generated monomeric proteoform framework, PRPH2 natively functions within the photoreceptor outer segment discs as a complex network of homodimers and PRPH2-ROM1 heterodimers.^19,101^ This multimeric architecture is stabilized by an intermolecular disulfide bridge at residue p.C150. Interestingly, our spatial analysis revealed that pathogenic variants driving pattern dystrophy (PD) significantly cluster around this specific interface, highlighting its structural vulnerability. Emerging structural evidence modeling higher-order PRPH2 complexes suggests that mutations at these interfaces alter the dimer interaction energy (ΔIE), disrupting physical assembly and modifying interaction boundaries without collapsing or destabilizing the core monomeric fold.^19,101^ This interface-specific pathology explains why mutations in these surface-exposed regions manifest as macular-dominant pattern dystrophies, whereas core-disrupting mutations lead to widespread RP. The correlation observed in our 3D map between the disruption of the highly basic, electropositive surface patch on the extracellular loop and localized macular dystrophy phenotypes suggests a distinct biochemical mechanism.^102^ Localized electrostatic surface potentials are well-established drivers of macromolecular docking, governing both protein-lipid interactions and the high-molecular-weight oligomeric assembly required for disk-rim morphogenesis.^93^ A charge inversion or alteration in this positive cluster likely disrupts crucial electrostatic interactions with negatively charged phospholipid headgroups (such as phosphatidylserine) or adjacent structural partners like ROM1.^78,103^ This hypothesis aligns with established biophysical models where precise positive surface charges dictate membrane-bending and macromolecular scaffolding, providing a compelling roadmap for future *in vitro* binding assays to trace the exact kinetic consequences of these missense variants.^103^

The AI-generated PRPH2 structural framework utilized in our pipeline agrees well with the experimentally determined PRPH2-ROM1 heterodimer complex (PDB ID: 7ZW1).^78^ Notably, while our framework highlights the dimerization interface as a hotspot for PD, there is currently no clinical or genetic evidence demonstrating that an isolated mutation of the p.C150 residue itself causes human retinal disease.^19^ This structural finding aligns with p.C150S knock-in mouse models showing that while this specific residue is not strictly mandatory for initial ROM1 binding, its disruption alters overall outer segment conformation, inducing a haploinsufficient state in rods and a dominant-negative effect in cones.^19^ Similarly, *ROM1* knock-out animals display a compensatory increase in PRPH2 disc content, and their subsequent delay in disc enclosure can be completely rescued via PRPH2 overexpression.^101^ Therefore, defects in p.C150-mediated heterodimerization alone are insufficient to trigger a disease phenotype; rather, the cumulative, multi-residue structural conformation and electrostatic interactions across the entire dimerization interface dictate the clinical outcome.^19^ Future population-scale studies tracking concurrent PRPH2 and ROM1 variants via WGS at the individual haploblock level will be required to definitively map these epistatic modifiers.

The distinct spatial segregation of clinical phenotypes observed across the 3D structure of PRPH2 aligns with an emerging paradigm in genomics, wherein the precise structural topology of a variant dictates its downstream molecular pathology, phenotypic manifestation, and inheritance pattern. A striking structural parallel is found in *KLHL7*-associated retinopathies, where pathogenic variants driving autosomal dominant retinitis pigmentosa (adRP) cluster exclusively within the internal core BTB and BACK domains, disrupting crucial E3-ubiquitin ligase complex assembly and inducing haploinsufficiency.^104^ In contrast, recessive alleles driving systemic Crisponi/Bohring-Opitz-like syndrome (BOS-3) segregate to the outer Kelch domain, preserving structural complex formation but modifying substrate specificity.^104^ Similarly, in RPGR, spatial clustering of missense variants within the regulator of chromosome condensation (RCC1)-like domain alters key macromolecular interactions with its ciliary partner, RPGRIP1. The precise coordinates and thermodynamic impact of these surface-exposed changes dictate the phenotypic divergence between classic X-linked RP and severe Leber congenital amaurosis (LCA).^105^ This principle extends beyond retinal disease. For example, missense variants mapped onto the three-dimensional structure of A-type nuclear lamin (encoded by *LMNA*) demonstrate a clear structure-phenotype correlation. Mutations mapping onto buried, stability-disrupting substitutions predominantly produce skeletal muscle disease, whereas surface-exposed substitutions clustering within distinct charged regions of the same domain instead produce lipodystrophy or premature-aging phenotypes.^106^ These examples reinforce that variant position and structural context, not gene identity or sequence conservation alone, are often the primary determinants of clinical outcome,

By contextualizing PRPH2 within this broader structural biology landscape, we demonstrate that 3D proteoform-phenotype mapping provides a robust, generalizable framework to guide domain-targeted therapeutic design. Gene therapy is now a reality for retinal dystrophy patients, but for poorly characterized mutations, as in PRPH2, designing appropriate clinical trials remains remarkably difficult. In patients with RP or rod-dominant phenotypes caused by rapidly degrading protein products, a gene supplementation approach is mandated to overcome haploinsufficiency. Conversely, classic gene supplementation would be highly ineffective for patients with cone-dominant, dominant-negative phenotypes stemming from stable, interface-disrupting mutant proteins that actively disrupt macromolecular assembly. Interpreting whether a variant causes a core thermodynamic folding collapse or an isolated surface-interface disruption is mandatory for the selection of precision clinical trial candidates.

Of the 20-to 30-thousand human genes, more than 3,500 are associated with documented genetic diseases.^107^ Traditional clinical genetic analysis successfully predicts pathogenicity for approximately 60% of identified variants, leaving a massive 40% categorized as VUS.^2^ This diagnostic gap is profoundly magnified in inherited retinal diseases, where over 72% of the roughly 16,000 reported variants remain classified as VUS, and the volume of experimentally determined 3D structures is vastly insufficient to inspect them.^21^ Approaches like the one presented here were simply not feasible before AlphaFold and AlphaMissense made high-confidence 3D structure prediction and large-scale sequence-based pathogenicity scoring available for proteins, including PRPH2, that had never been experimentally solved. This capability is transformative, particularly for the many disease-relevant proteins that still lack a solved structure, yet it remains underutilized across the broader genetics community relative to its potential. To overcome this systemic bottleneck, our approach takes advantage of high-fidelity, AI-based structure predictions, which, when coupled with human expert adjudication bridging basic and clinical science, successfully decodes the phenotypic trajectories of uncharacterized mutations. To establish a definitive baseline framework and prevent structural signal dilution by clinical pleiotropy, this study intentionally focused on variants associated with singular, well-defined clinical phenotypes. While isolating this high-fidelity baseline dataset was a prerequisite, the underlying 3D proteoform-phenotype workflow is a generalizable blueprint that can be applied to other complex loci where the same therapeutic stakes demand mechanistic, per-variant resolution rather than genome-wide triage. Demonstrating that spatial mapping resolves the severe pleiotropy of PRPH2, a gene long considered intractable to genotype-phenotype correlation, establishes that this depth-focused approach can be extended, one therapeutically actionable locus at a time, to other multi-phenotypic Mendelian diseases.

The data dependencies of this spatial-mapping strategy align with two ongoing shifts in biomedical data availability, not as engines for proteome-scale automation, but as complementary inputs that still require expert structural and clinical adjudication. First, translational genetics is transitioning from isolated case reports toward deeply phenotyped national biobanks and international registries, such as the UK Biobank, the All of Us Research Program, and specialized consortia like the International Inherited Retinal Dystrophy Consortium.^108,109^ These resources supply the standardized, imaging-backed phenotype catalogs that make confident structure-phenotype mapping possible for a given disease gene. Second, the historical bottleneck of low-throughput biochemistry is being addressed by high-throughput functional proteomics: multiplexed assays such as Deep Mutational Scanning (DMS) and Variant Effect Mapping (VEM) now generate structure-wide experimental measurements of the thermodynamic and functional penalties of thousands of variants in parallel. Critically, neither data stream removes the need for human-in-the-loop interpretation; rather, each enriches the evidence that expert adjudicators integrate with AI-generated structural models. By using these models as a spatial scaffold, the framework introduced here provides a reproducible template for fusing emerging biobank and proteomics data into mechanism-resolved variant calls. In an era of expanding precision medicine that includes antisense oligonucleotides (ASOs), gene augmentation, and base editing, this depth-oriented analysis offers a powerful, complementary modality to genome-scale predictors, bridging the chasm between variants of uncertain significance and the mechanistically stratified patient cohorts that small, high-stakes clinical trials require.

## Materials and Methods

### Ethical approval

Retrospective analysis of patients with documented mutations in *PRPH2* was performed. Patients were evaluated either at the Byers Eye Institute at Stanford University or at the Edward S. Harkness Eye Institute at Columbia University Medical Center. Patient consent was obtained as per Stanford University Institutional Review Board-approved protocols IRB-42554, IRB-42754, or Columbia University Institutional Review Board-approved protocol IRB-AAAB6560. All procedures were reviewed and in accordance with the tenets of the Declaration of Helsinki. The data presented in this study was procured through retrospective chart review and was not identifiable to any individual patient.

### Phenotypic ascertainment

Patients were seen by retinal specialists (VBM and SHT) at two tertiary academic eye centers. They underwent genetic sequencing (were screened for *PRPH2* variants), and thorough examination by a retinal physician, i.e., fundus photography with an FF 450plus Fundus Camera (Carl Zeiss Meditec AG, Germany and Optos California ICG, Scotland). FAF images via confocal scanning-laser ophthalmoscope (Heidelberg Retina Angiograph 2, Heidelberg Engineering, Germany) and fundus illuminated with an argon laser light (488 nm) and fluorescence assessed via band pass filter with a short wavelength cutoff (495 nm). Simultaneous FAF and spectral domain optical coherence tomography (SD-OCT) images were acquired via Spectralis HRA+OCT (Heidelberg Engineering, Germany).

### Patient sequencing

Sequencing was via Invitae Inherited Retinal Disorder, the Blueprint Genetics My Retina Tracker panel, Retinal Dystrophy, or MVL Vision panels. For some, whole-exome sequencing replaced candidate gene testing. Variants were annotated using the ClinVar database and the literature and classified as novel if no publication discussed that variant, or VUS if previously reported but no sufficient data to declare its pathogenicity.

### Establishment of the PRPH2 variant database

To construct a high-confidence, structurally actionable database for PRPH2 topological mapping, a multi-layered, stringent multi-step exclusion cascade was executed on raw genetic variants harvested from the ClinVar repository (n = 503 initial *PRPH2* entries; accessed April 11, 2022).^21^ Entries lacking rigorous evidence packages, specifically those designated with a 0-star review status (e.g., “no assertion criteria provided” or “no assertion provided”), were systematically discarded. The remaining variants were passed through a molecular consequence filter to restrict the cohort to variants causing direct translational or coding changes. Variants residing in non-coding zones, including untranslated regions, deep intronic segments altering non-canonical splice donor/acceptor sites, flanking regions, and non-coding RNAs, were removed. To bridge the translational gap between genomic variation and clinical manifestation, the variants were subjected to rigorous phenotypic curation. To ensure a high level of clinical stringency, variants were not selected solely based on repository annotations. Instead, variant classifications and their associated clinical trajectories were cross-referenced and verified against peer-reviewed, published case reports containing explicit clinical phenotyping and diagnostic structural imaging, including OCT, FAF, and/or ERG.^5,22–75^ The decision to exclude the highly abundant Cone Dystrophy (CD) and Cone-Rod Dystrophy (CRD) variants during the initial calibration phase was a deliberate signal-to-noise optimization strategy. By defining the absolute spatial properties of clear, non-overlapping polar clinical presentations first (RP vs. CACD), this framework provides the baseline scaffold needed to systematically deconvolve intermediate spectrum phenotypes in future studies. Through this systematic review of the literature and public repositories, a comprehensive dataset of 164 verified PRPH2 variants was compiled. To validate our baseline variant classifications against current clinical consensus, a cross-comparison was performed against the ClinVar database in June 2026. Variants were evaluated for exact genomic and protein matches. While minor classification shifts (e.g., shifts to uncertain significance or emerging conflicting classifications) were observed for a small subset of variants due to evolving laboratory submissions, these updates did not significantly impact the overall cohort trends or the primary conclusions of our downstream analyses (Supplemental Table 2). To supplement and validate the repository-derived data, we integrated an independent, institutional dataset comprising 31 deep-phenotyped inherited retinal dystrophy probands harboring mutations in PRPH2, accounting for 23 unique mutation events across the independent patient cohort. To ensure uniform database cross-referencing, raw genomic variants tracked across conflicting assemblies (GRCh37 and GRCh38) were harmonized to standard GRCh37 coordinates via a site-specific liftover correction before population-scale evaluation. The curated variants were subsequently evaluated against population-scale tracks via an automated pipeline querying the Genome Aggregation Database (gnomAD v4 global tracks). To filter out common, benign background polymorphisms, variants with AF > 0.0005 were excluded. Following this population-scale filter, all remaining clinically curated, disease-driving variants were either entirely absent from population tracks or returned an exomic/genomic AF of 0, confirming their population rarity and high-penetrance potential. To finalize the baseline required for position-specific structural modeling, these unique variants were passed through a final structural capability filter. Truncating, premature termination mutations, specifically nonsense alterations (“stop gained” mechanisms), frameshifts, and in-frame indels, were systematically segregated into a clinical comparative matrix (n = 66). While clinically deleterious, these variants trigger nonsense-mediated decay or result in total macro-structural deletions, rendering them incompatible with position-specific 3D atomic mapping. The remaining missense alterations were partitioned into an unclassified background track comprising variants of unknown significance (VUS; n = 52) and a final, high-confidence 3D Phenotype-Mapping Calibration Baseline of 46 clinically confirmed, imaging-backed pathogenic single-nucleotide missense substitutions (**Extended Data Table 1**). These calibrated variants were projected directly onto the three-dimensional atomic coordinates of the PRPH2 proteoform matrix. Local physical microenvironments were computationally resolved by calculating site-directed thermodynamic stability changes (ΔΔG), relative solvent accessibility (RSA), and local alpha-helical structural topology, providing an empirical spatial blueprint to resolve phenotypic bifurcation based on structural microenvironment constraints.

### Structural modeling and in silico calculations

The AlphaFold database (version 2.0.0; AlphaFold ID: AF-P23942-F1) provided the PRPH2 structure model.^9^ To establish biologically meaningful phenotypic zones, variants were not clustered using arbitrary geometric boundaries (e.g., spatial spheres or cubes), which fail to capture asymmetric protein dynamics. Instead, clusters were defined by shared quantitative biophysical metrics. Biophysical metrics for each missense variant were computed utilizing the Site Directed Mutator (SDM) server, which employs an environment-specific substitution scoring matrix to calculate the thermodynamic consequences of amino acid substitutions (i.e., ΔΔGfold) on the PRPH2 protein structure.^82^ Relative solvent accessibility (RSA) was expressed as a percentage (0–100%) representing the exposure of the residue side chain to the aqueous solvent matrix. Residual positions with an RSA < 15% were defined as structurally buried or interior core loci, while positions with an RSA > 25% were designated as surface-exposed. Atomic depth was measured in Å as the distance from the residue’s geometric center to the nearest solvent-accessible surface atom. The predicted change in free energy (measured in kcal/mol) associated with mutation-induced protein stability shifts. Highly negative values (ΔΔG < −2.0 kcal/mol) were classified as severely destabilizing to tertiary fold architecture. Electrostatic surface potential calculations were performed using the Adaptive Poisson-Boltzmann Solver (APBS).^110^ The unmutated and mutant structural models were first prepared using PDB2PQR software to assign charge and radius parameters based on the PARSE force field, reconstruct missing heavy atoms, and optimize hydrogen-bonding networks at a simulated physiological pH of 7.4.^111^ Continuum electrostatics calculations were subsequently executed in APBS by solving the linearized Poisson-Boltzmann equation. The spatial charge distribution and solvent-accessible surface potentials were mapped onto a three-dimensional grid using a dielectric constant of 78.5.4 for the aqueous solvent and 2.0 for the hydrophobic protein core. Localized electrostatic potential shifts at variant coordinate points were quantified in units of dimensionless thermal energy. Deep learning-based pathogenic likelihoods and categorical classifications for all *PRPH2* missense variants and single-amino-acid substitutions were acquired directly from the AlphaMissense repository via the Google Cloud Storage framework (dm_alphamissense).^12^ To incorporate integrated ensemble-based scoring, we utilized MetaRNN and BayesDel frameworks.^76,77^ MetaRNN scores were extracted to capture context-dependent nucleotide and amino-acid conservation features utilizing a deep recurrent neural network architecture optimized to integrate high-throughput sequencing constraints and flanking genomic contexts.^76^ BayesDel predictions were cross-referenced as an ensemble meta-score that integrates functional genomic data, computational predictions, and multi-species alignments using a Bayesian framework without clinical database over-fitting bias.^77^ All data wrangling and vector alignments were performed in R Studio (version 2025.09.1+401, R version 4.5.1) utilizing the tidyverse framework (v2.0.0) and compiled into comprehensive reference matrices using explicit relational left-joins.^112^ Continuous biophysical data (ΔΔG and RSA percentage) between VUS and pathogenic groups were analyzed via independent two-tailed Mann-Whitney U tests (Kruskal-Wallis with post-hoc corrections for multi-group clinical phenotype comparisons). All statistical testing and multi-panel scatter visualizations were built and rendered using GraphPad Prism 11. Figures were generated using Pymol (The PyMOL Molecular Graphics System, Version 2.0 Schrödinger, LLC).

### Human-in-the-loop AI (HITL-AI) variant adjudication

Variant interpretation followed a structured human-in-the-loop workflow rather than a purely automated pipeline. Each variant was first mapped onto the AI-generated PRPH2 structural scaffold, and its predicted structural perturbation was evaluated using predicted change in protein stability (ΔΔG), position relative to known functional domains (e.g., the D2 intradiscal loop), and predicted disruption of the residue’s disulfide network and relevant adjacent structural interfaces. All variants were then reviewed by multiple domain experts among the authors, without masking. For each variant, reviewers assessed the predicted structural change in the context of (1) its location relative to known functional domains, (2) historically validated structure-function relationships, and (3) the corresponding multimodal clinical imaging phenotype. Reviewers classified each variant’s structural consequence and its consistency with the observed phenotype. Final structure-phenotype assignments were reached by consensus among the reviewing experts, and all variant classifications and VUS prioritizations reported here reflect this expert consensus rather than raw model output.

## Data Availability

All data produced in the present study are available upon reasonable request to the authors. PRPH2 variant information used in this study can be obtained from the publicly accessible ClinVar by applying prph2[gene]
search (https://www.ncbi.nlm.nih.gov/clinvar/). PRPH2 AlphaFold protein structure model used in this study is available from the publicly accessible AlphaFold Protein Structure Database, accession number P23942
(https://alphafold.ebi.ac.uk/entry/P23942).

## Data Availability

PRPH2 variant information used in this study can be obtained from the publicly accessible ClinVar by applying prph2[gene] search (https://www.ncbi.nlm.nih.gov/clinvar/). PRPH2 AlphaFold protein structure model used in this study is available from the publicly accessible AlphaFold Protein Structure Database, accession number P23942 (https://alphafold.ebi.ac.uk/entry/P23942).

## Code Availability

No new code or algorithm was generated or used.

## Credit Authorship Contribution Statement

All authors attest that they meet the current ICMJE criteria for Authorship. Dr. Mahajan had full access to all the data in the study and takes responsibility for the integrity of the data and the accuracy of the data analysis. Study concept and design: YJS, AGB, VBM. Acquisition of data: YJS, ASL, GV, AK, CP, JKO, MGSC, SHT. Analysis and interpretation of data: YJS, ASL, GV, AK, SHT, JTV, CP, DKR, VBM. Drafting of the manuscript: YJS, GV, ASL, AK, JTV, CP, DKR, VBM. Critical revision of the manuscript for important intellectual content: YJS, GV, YS, AGB, SHT, VBM. Obtained funding: VBM. Administrative, technical, and material support: YJS, AK, JKO, SHT, VBM. Study supervision: VBM.

## Institutional Review Board Statement

Patient consent was obtained as per Stanford University Institutional Review Board-approved protocols IRB-42554, IRB-42754, or Columbia University Institutional Review Board-approved protocol IRB-AAAB6560.

## Supplementary Materials

**Supplemental Figure 1.**
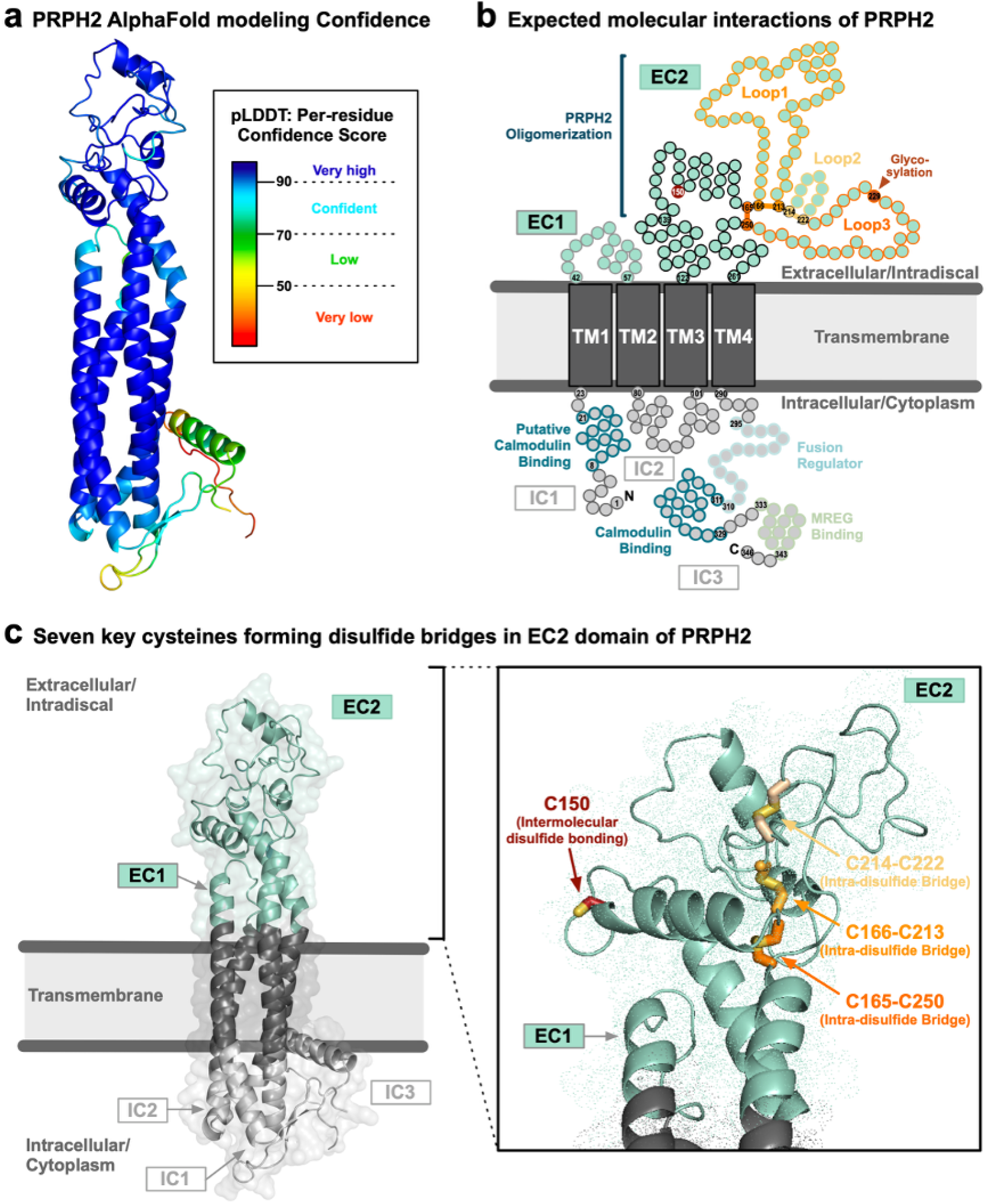
Quality assessment of PRPH2 AlphaFold model. a, Modelling confidence of PRPH2 AlphaFold model (AlphaFold ID: AF-P23942-F1) shows that overall PRPH2 AlphaFold model is predicted in very high confidence except the extreme N-termini and C-termini. The predicted structure of residues with per-residue confidence score (pLDDT) greater than 90 are very high in confidence (colored in deep blue). Residues with pLDDT between 70 and 90 are considered to be confidently modelled (colored in cyan). Residues with pLDDT between 50 to 70 are low in confidence. Residues with pLDDT lower than 50 are very low in modeling confidence. b, 2D domain map with anticipated domains and key molecular interactions based on prior studies. The three intramolecular disulfide bridges form three distinctive loops of EC2 domain. c, PRPH2 AlphaFold model properly modelled the three intramolecular disulfide bridges forming EC2 loops. Cys150, intermolecular disulfide bridging cysteine, is also modeled at the solvent exposed area as expected.

**Supplemental Figure 2.**
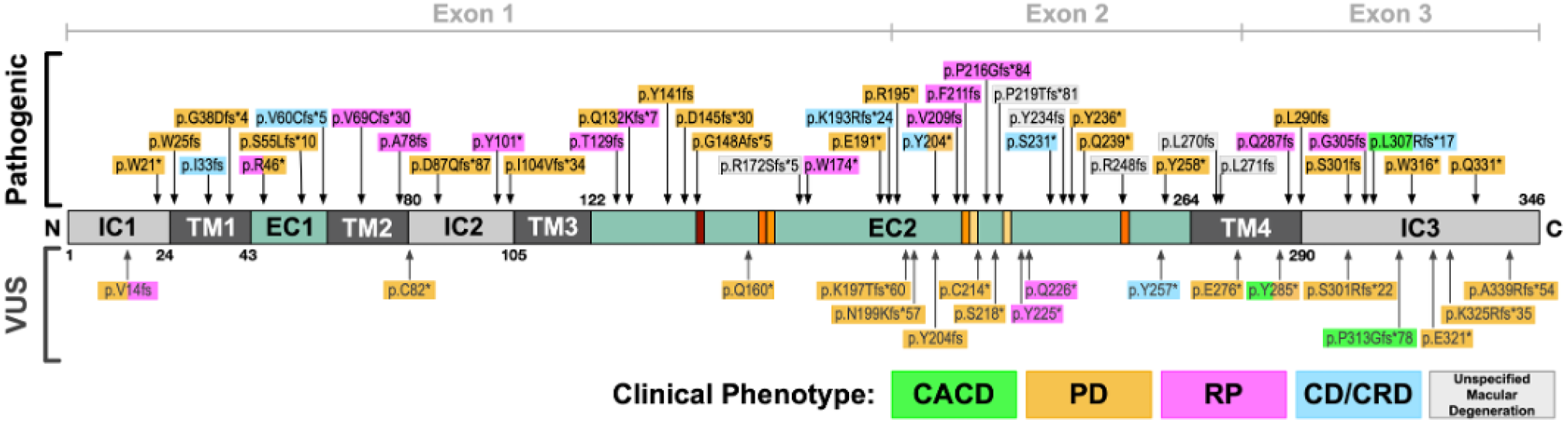
Linear mapping of nonsense and frameshift PRPH2 variants. Previously reported PRPH2 nonsense and frameshift variants are mapped to a linear schematic (primary structure map; domain topology map) of the PRPH2 protein. Pathogenic variants and VUS are shown above and under the map, respectively. Variants shown in orange, magenta, green, cyan, and gray boxes were identified in the patients displaying PD, RP, CACD, CD/CRD, and unspecified macular degeneration, respectively.

**Supplemental Figure 3.**
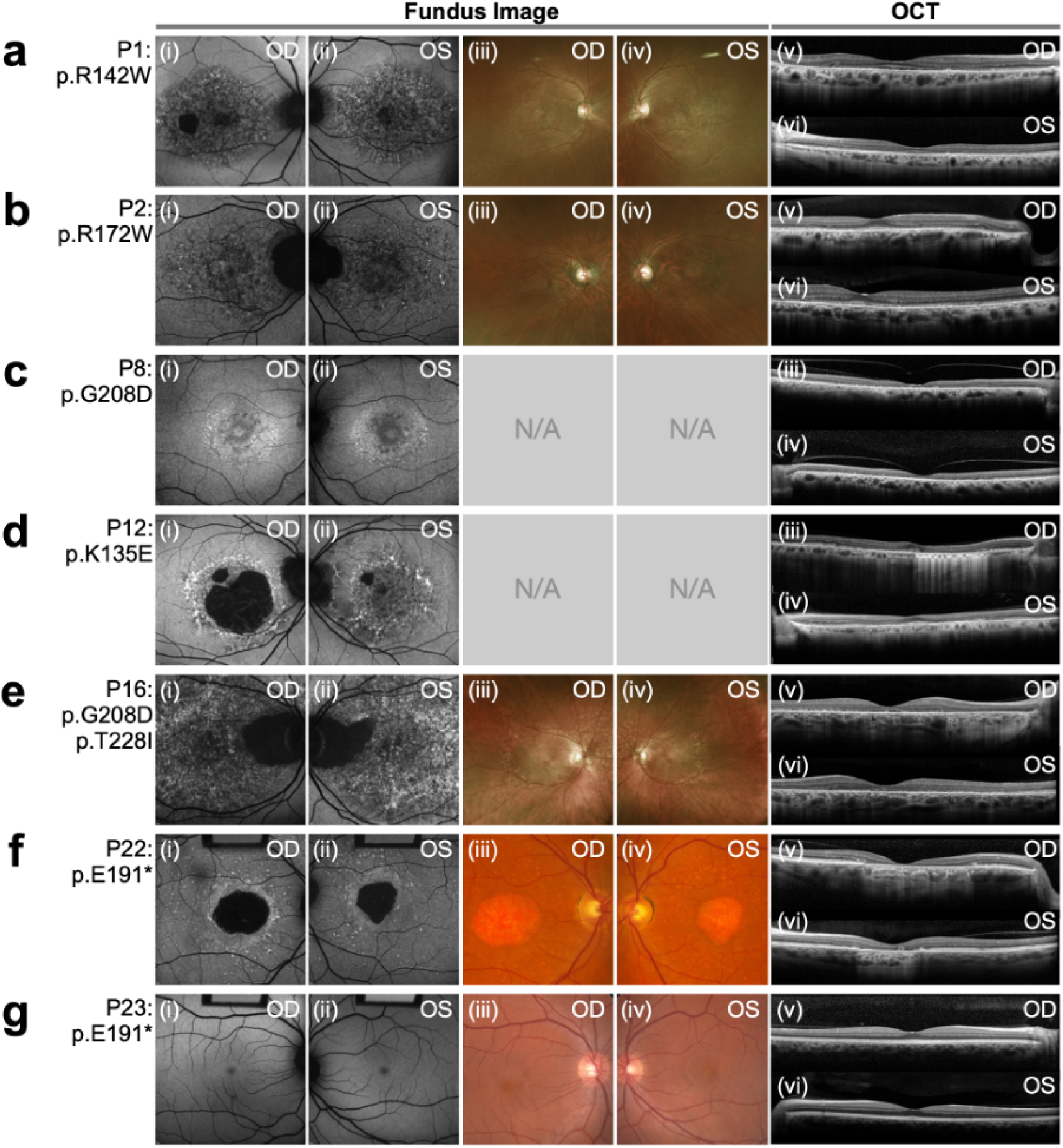
Clinical features of patients with central areolar choroidal dystrophy (CACD). a, P1 (p.R142W) displays classic signs of CACD with a speckled fundus autofluorescence (FAF) pattern and granular central macular atrophy with punctate pale yellow lesions OU. b, Clinical imaging of P2 (p.R172W) shows the classic CACD phenotype including speckled pattern of hypofluorescence OU, agranular appearance of the macula along with some peripapillary atrophy, and slight loss of outer retinal structure on OCT. c, FAF of P8 (p.G208D and VUS p.T228I) shows an area of classically speckled AF with peripapillary involvement. Wide-angle fundus photography reveals diffuse yellow deposits in the posterior pole that were absent on FAF. OCT shows increased hyperreflectivity in the outer retinal area. d, FAF for Patient 12 (P12; p.K135E) shows a well-demarcated area of decreased autofluorescence along with a prominent paracentral hyperfluorescent ring OD and a speckled pattern of AF OS. OCT shows bilateral loss of outer retinal layers and disorganization of retinal layers OU along with hyperfluorescent lesions in the retinal pigment epithelium (RPE) OD. e, FAF of P16 (p.G208D) reveals a bullseye pattern of atrophy with refractile crystals and macular atrophy on OCT. f, FAF, fundus photography, and OCT of P22 (p.E191*) shows well-demarcated area of central macular atrophy bilaterally along with a paracentral hyperfluorescent ring OD and hyperfluorescent flecks OU. g, FAF, fundus photography, and OCT for P23 (also with p.E191*) are unremarkable.

**Supplemental Figure 4.**
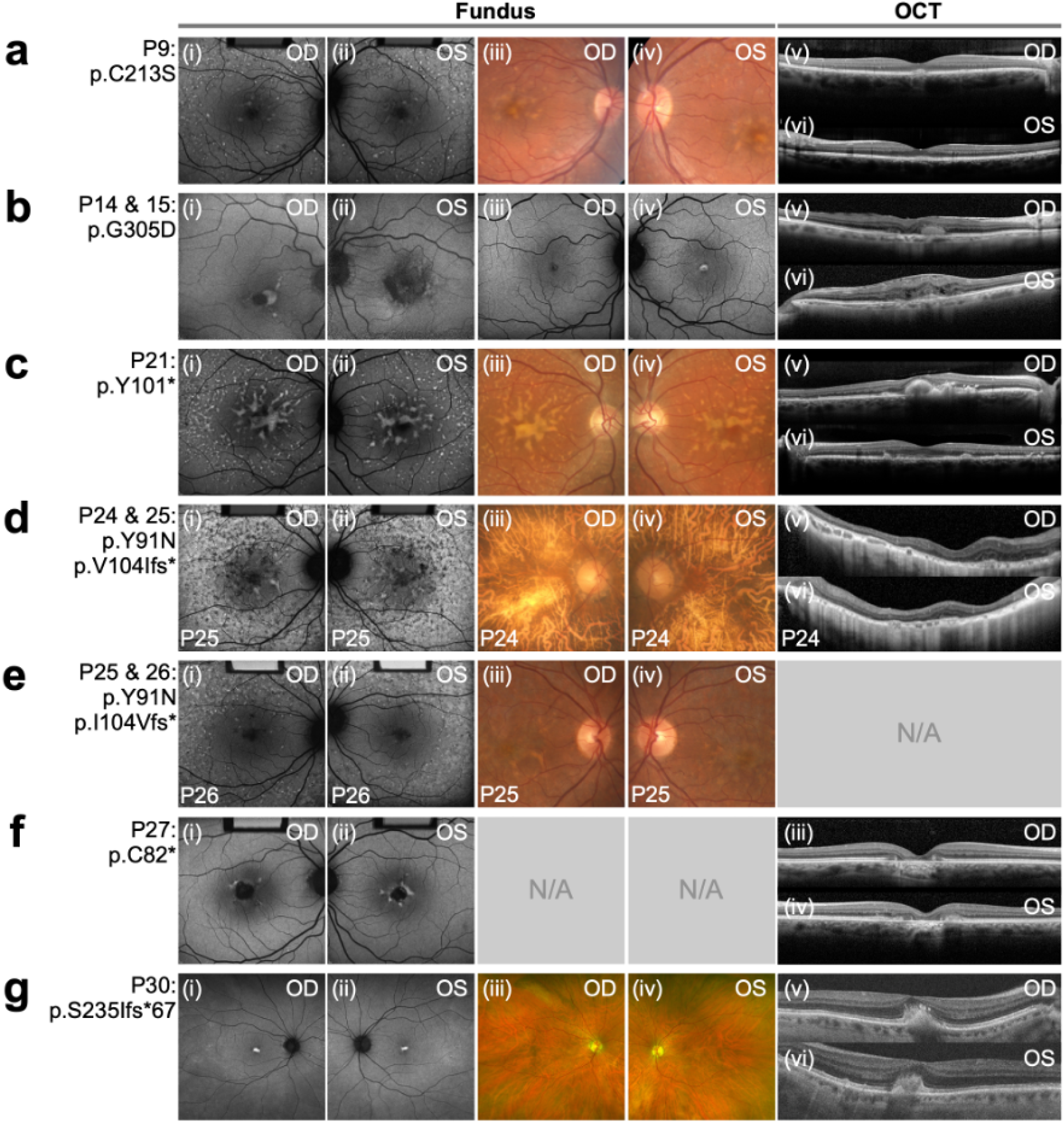
Clinical features of patients with pattern dystrophy (PD). a, FAF of P9 (p.C213S) displayed bilateral fleck-like lesions and yellow deposits in the macula were seen in the fundus images. OCT scan of the right eye showed drusen and mostly intact retinal layers. b, FAF OD (i) of P14 (p.G305D) demonstrates a well-demarcated, hyperfluorescent lesion in the central macula. FAF OS (ii) shows diffusely mottled hypofluorescence in the left macula. OCT OD (v) of P14 shows the location of the vitelliform material in the subretinal space, nasal to the right fovea. OCT OS (vi) highlights subretinal fibrosis and disorganization of retinal layers directly surrounding the fovea. FAF OU (iii-iv) of P15 (p.G305D) shows a small, hyperfluorescent vitelliform-like lesion in the left fovea. c, Fundus photography of P21 (p.Y101*) showed extensive pisciform lesions which are all hyperfluorescent on AF. Bilateral OCT through the yellow patches shows large hyperfluorescent subretinal, drusen-like deposits involving the fovea. d and e, P24 (p.Y91N and p.I104V*) shows end-stage retinal atrophy phenocopying Stargardt Disease. Fundus photography for P25 and P26 with the same variants displayed bilateral hyperfluorescent flecks around the macula. f, FAF of P27 (p.C82*) showed butterfly-shaped hyperfluorescent lesions surrounding a small area of central RPE atrophy that is more notable OS than OD. Bilateral OCT confirmed deepening of the fovea. g, FAF of P30 (p.S235Ifs*67) shows hyperfluorescent lesions in the fovea OU that are located in the subretinal space as seen on OCT. There are also small, scattered hypofluorescent spots in the periphery OU.

**Supplemental Figure 5.**
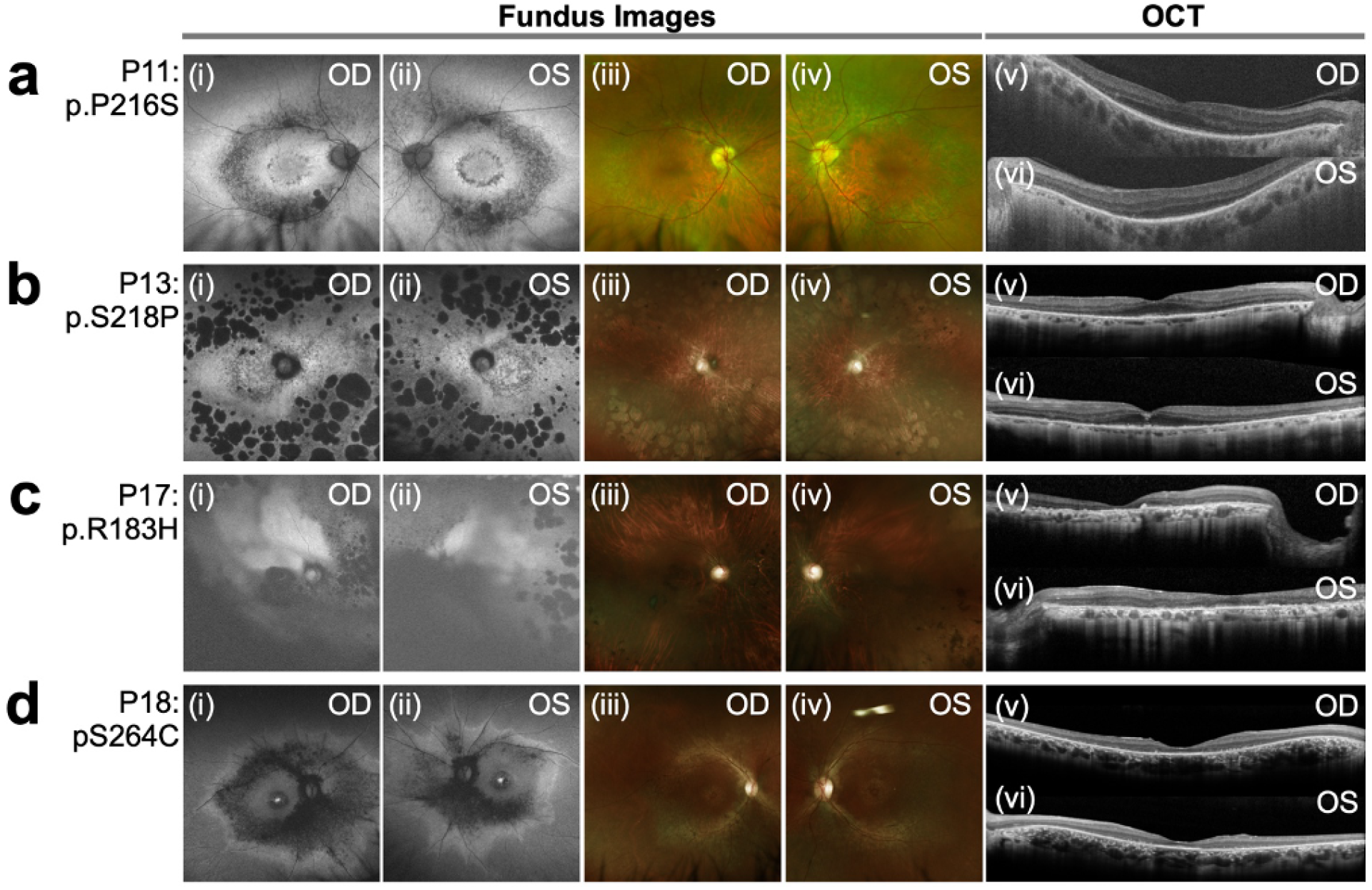
Clinical features of patients with retinitis pigmentosa (RP). a, Wide-angled fundus photography, FAF and OCT scan of Patient 11 (P11; p.P216S) showed granular changes along the posterior pole with a distinct ring of atrophy around the macula corresponding to thinning on OCT. b, FAF for P13 (p.S218P) shows granular appearance to the macula with numerous lacunar lesions in the posterior pole and intraretinal pigment migration. c, Wide-angled fundus photography, FAF and OCT scan of P17 (p.R183H) revealed widespread retinal atrophy with superior lacunar degeneration and pigmentary changes in the periphery. d, FAF of P18 (p.S264C) showed granular appearance to the macula with several hypofluorescent lacunar lesions in the posterior pole. OCT scan showed RPE loss.

**Supplemental Figure 6.**
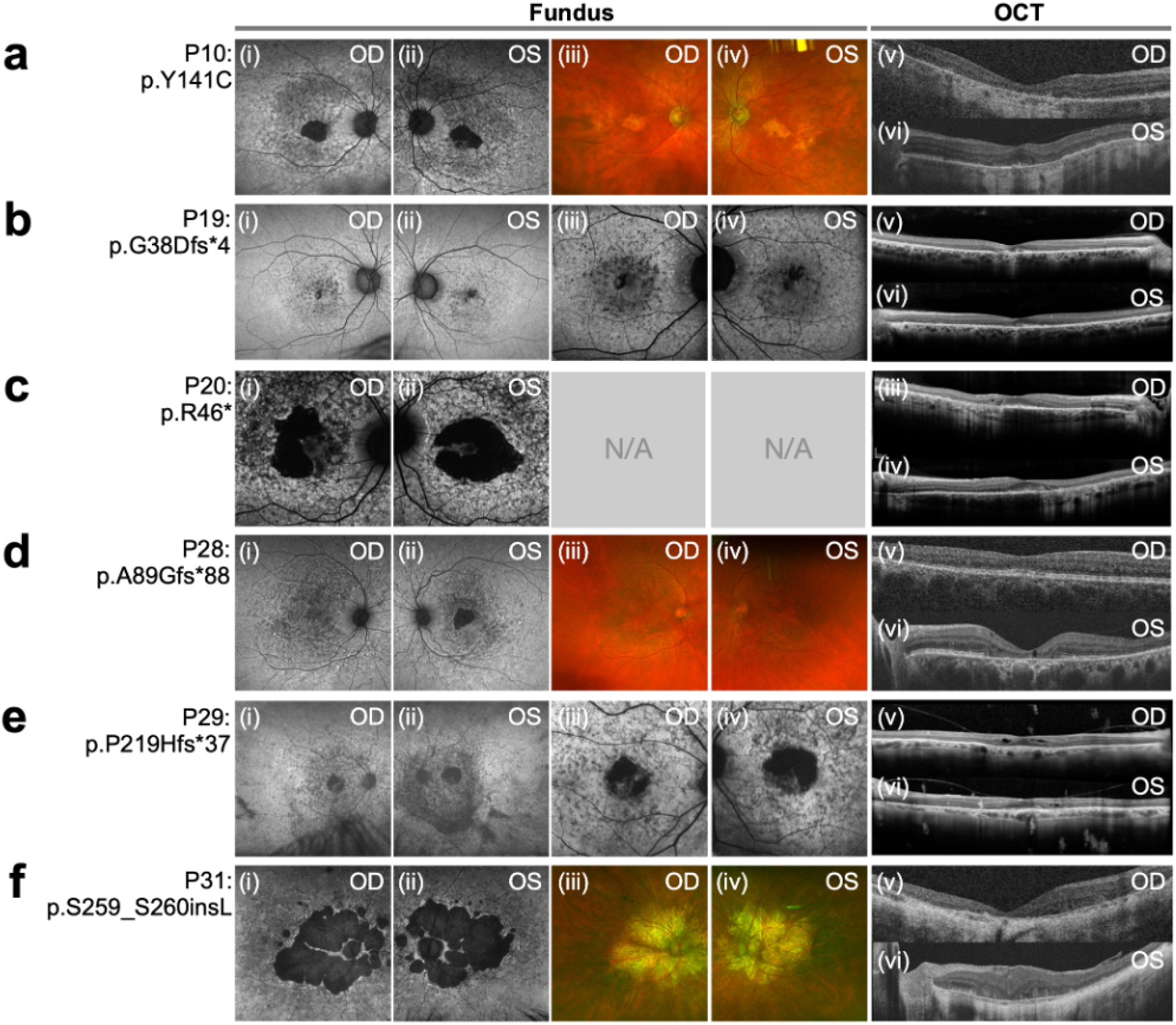
Clinical features of patients with cone/cone-rod dystrophy (CD/CRD). a, FAF of P10 (p.Y141C) shows an area of atrophy at the fovea. peripapillary sparing on FAF. OCT scan showed irregular contour, EZ and RPE loss seen in OCT. b, FAF of Patient 19 (P19; p.G38Dfs*4) showed bilateral speckled hypo-autofluorescence that is limited to the central macula. OCT scan revealed some loss of the outer nuclear layer. c, FAF of P20 (p.R46*) showed a well-demarcated area of central macular atrophy with diffuse hypofluorescent specks. OCT scan showed disorganization of retinal layers OU. d, Wide-angle fundus photography and FAF of P28 (p.A89Gfs*88) showed macular pigmentary changes, drusen, granularity at fovea OU, and a pisciform-like lesion in the periphery OU. OCT scan showed distortion of the RPE OS. e, FAF of P29 (p.P219Hfs*37) showed paracentral lesion with jagged border of hypofluorescence around the macula OU. OCT showed intraretinal cysts and partial posterior vitreous detachment (PVD). f, FAF and wide-angle fundus photography of P31 (p.S259_S260insL) revealed severe, diffuse retinal atrophy. Partial peripapillary sparing on FAF. OCT scan showed distortion of the RPE OU.

**Supplemental Figure 7.**
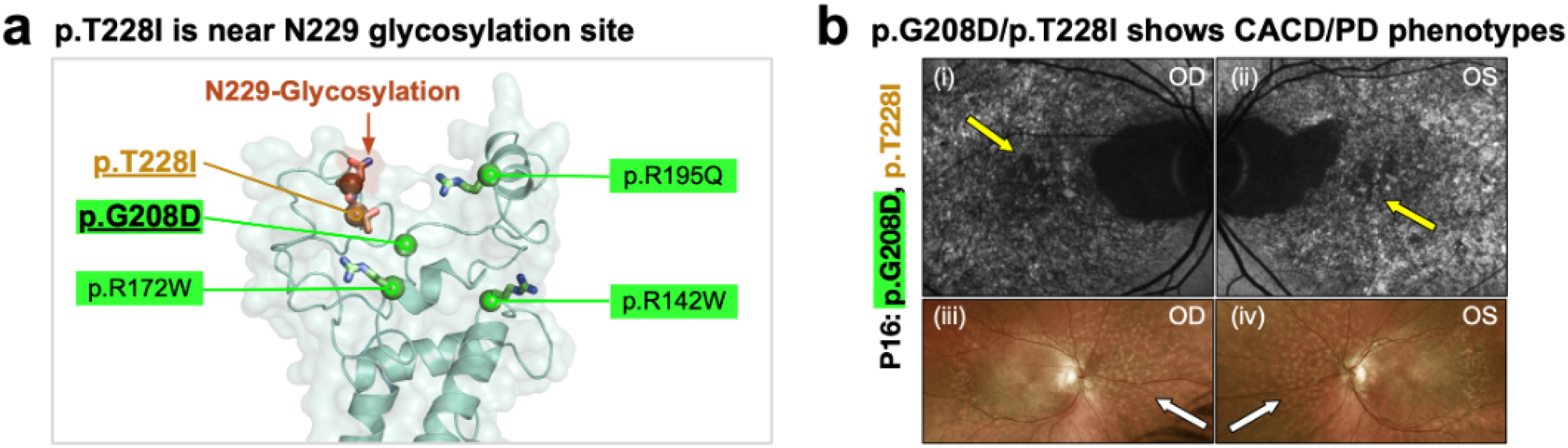
Proteotype-phenotype analysis of a patient with two different PRPH2 variants. a, p.T228I, previously reported in PD patients (light brown residue), was right by the Asn229 glycosylation site (brown residue). b, Patient 16 (P16) harboring p.G208D (pathogenic CACD variant) and p.T228I displayed combined phenotypes of CACD and PD. This is illustrated by fundus autofluorescence images (i and ii) showing the speckled pattern of hyper and hypo-autofluorescence typical in CACD, but widefield fundus photographs (iii and iv) showing the yellow fleck-like lesions (white arrows) in the posterior pole typical in STG-PD.

**Supplemental Figure 8.**
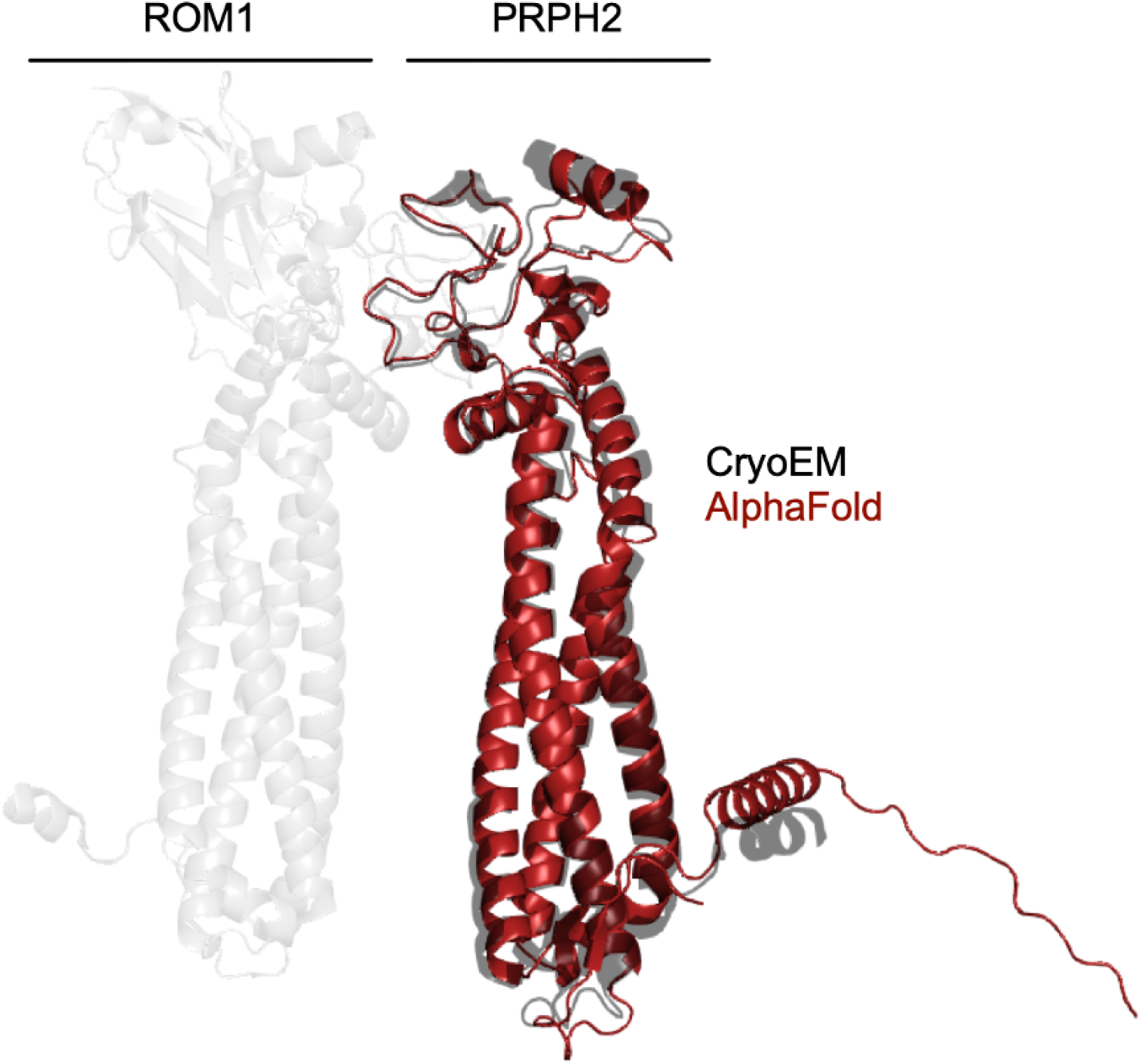
Comparison of PRPH2 AlphaFold model to cryogenic electron microscopy structure. Cryogenic electron microscopy structure of PRPH2 in complex with ROM1 validates AlphaFold model. Cα RMSD = 1.397 (2122 to 2122 atoms.)

**Supplemental Figure 9.**
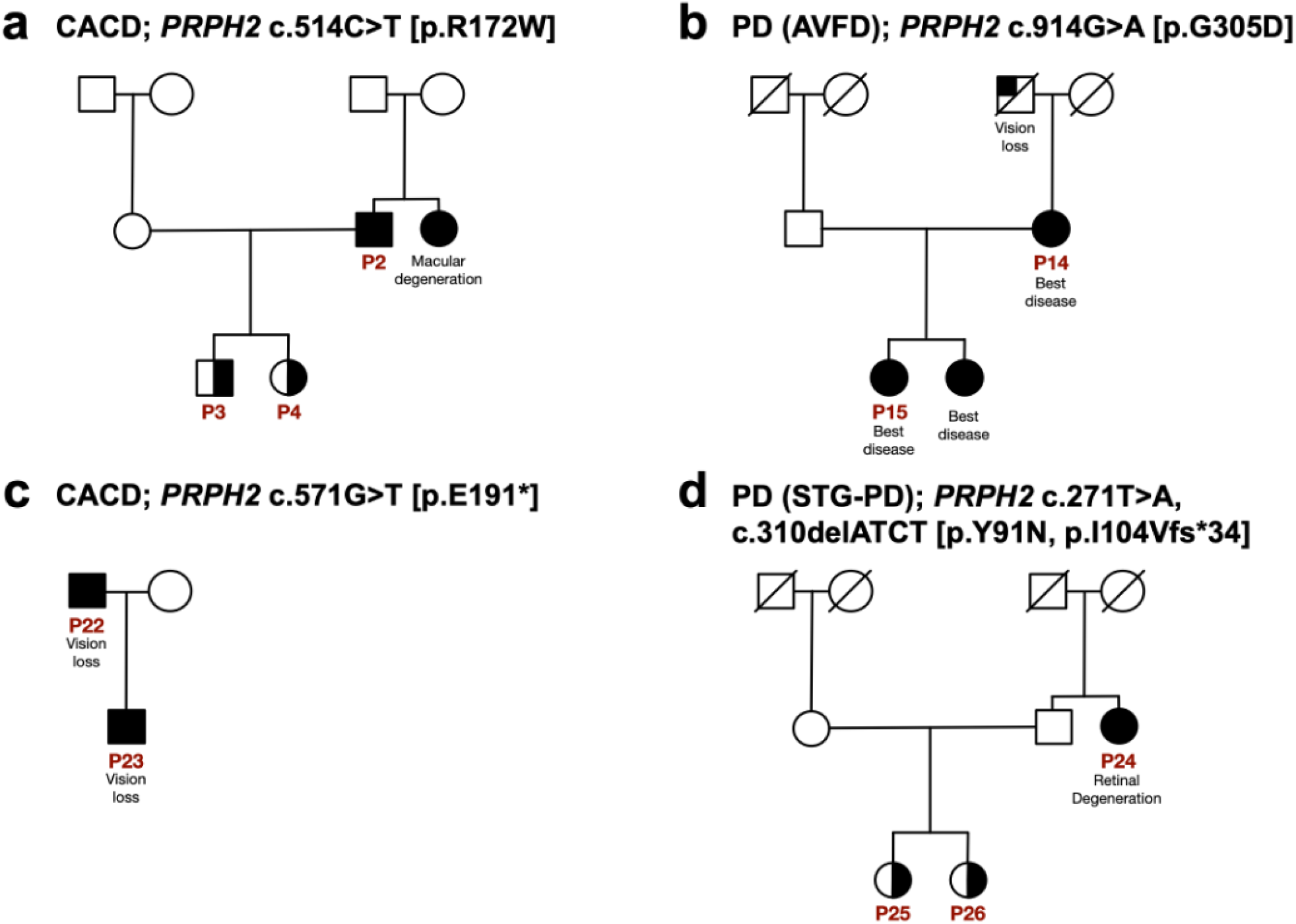
Pedigrees of related patients with PRPH2 variants. Multi-generational pedigrees display patterns of inheritance of PRPH2 variants within the same family. Circles represent females, and squares represent males. Symbols with a diagonal line indicate that the person is deceased. Filled symbols indicate that the person is affected with a, PRPH2 c.514C>T [p.R172W], b, PRPH2 c.914G>A [p.G305D], c, PRPH2 c.571G>T [p.E191*], or d, PRPH2 c.271T>A, c.310delATCT [p.Y91N, p.I104Vfs*34].

**Supplemental Table 1.**
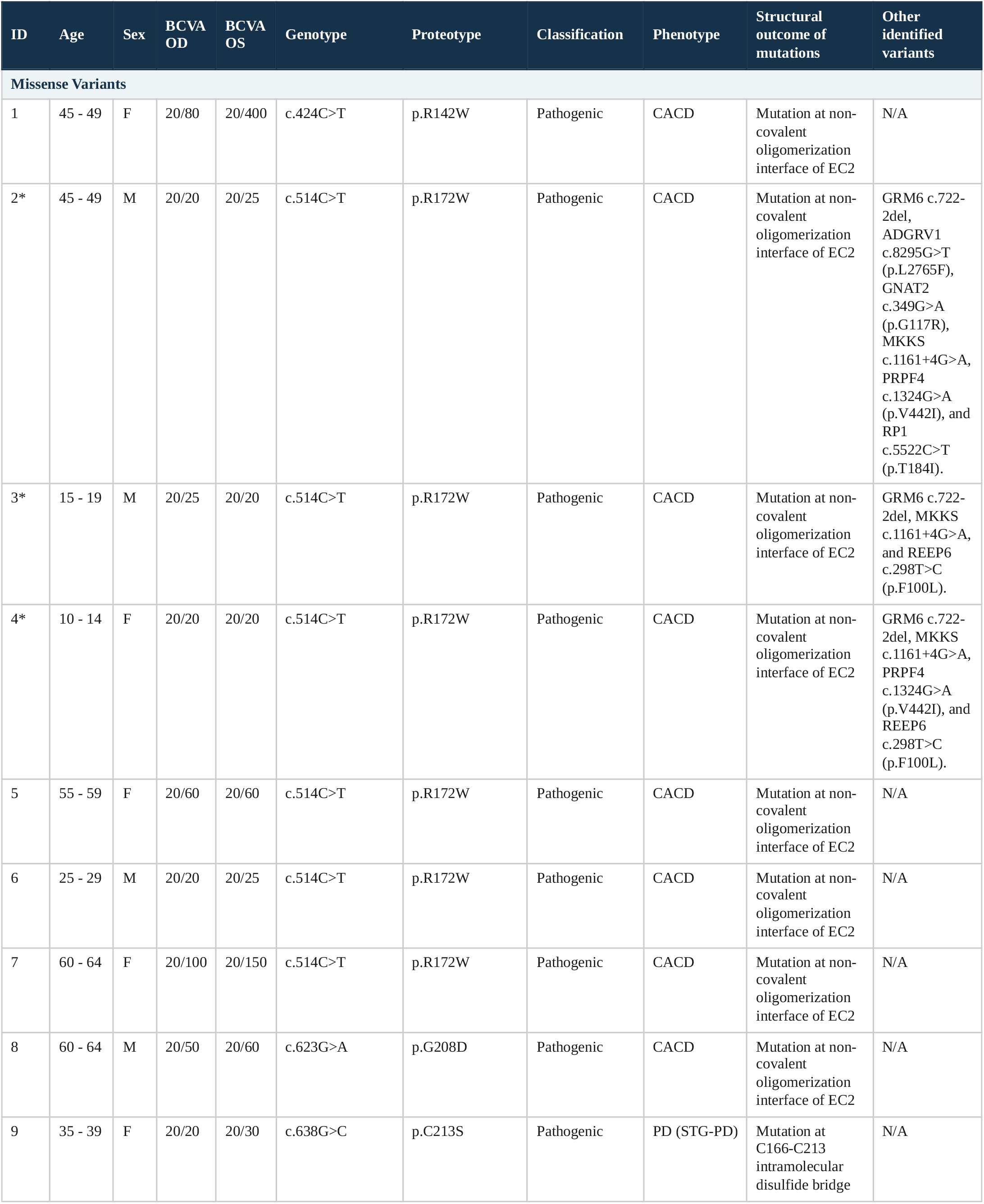
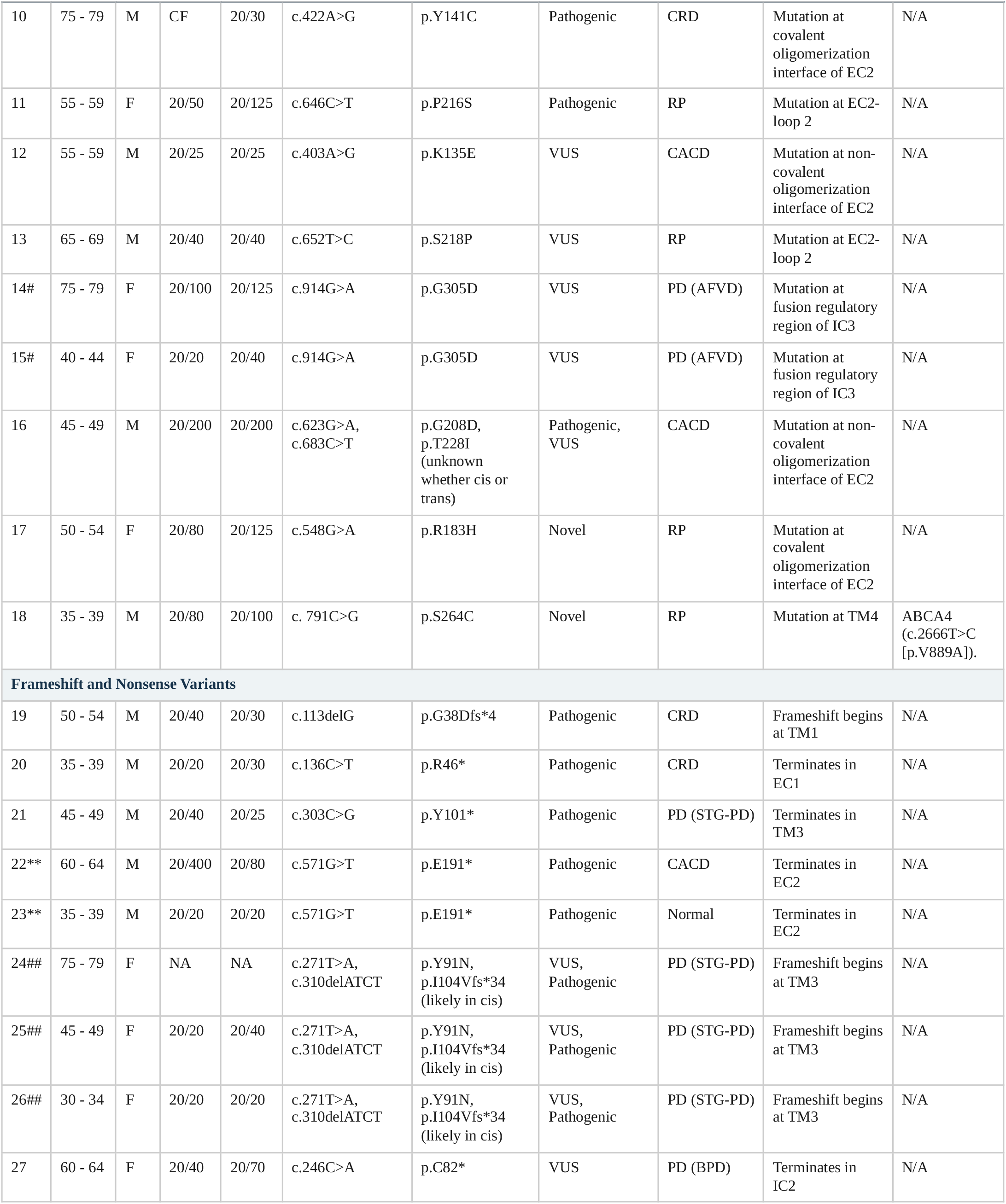
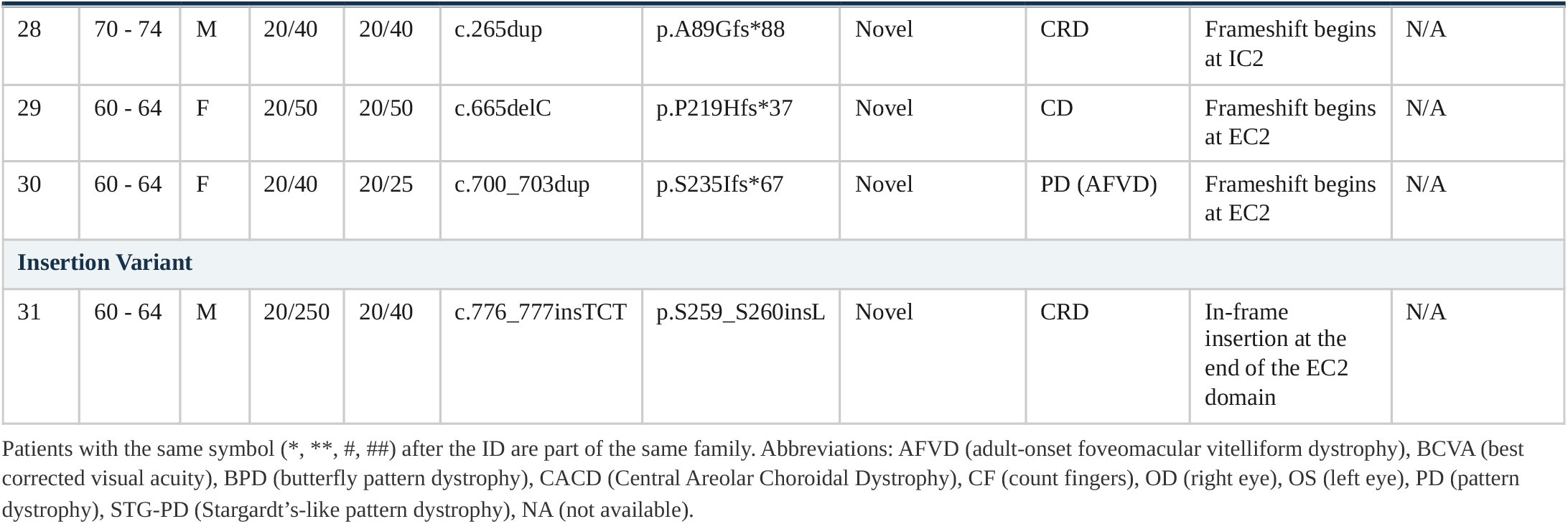
Summary of genotype, proteotype, and phenotype in 31 patients with PRPH2 variants.

| ID | Age | Sex | BCVA<br>OD | BCVA<br>OS | Genotype | Proteotype | Classification | Phenotype | Structural<br>outcome of<br>mutations | Other<br>identified<br>variants |
| --- | --- | --- | --- | --- | --- | --- | --- | --- | --- | --- |
| **Missense Variants** |  |  |  |  |  |  |  |  |  |  |
| 1 | 45 - 49 | F | 20/80 | 20/400 | c.424C>T | p.R142W | Pathogenic | CACD | Mutation at non-covalent oligomerization interface of EC2 | N/A |
| 2* | 45 - 49 | M | 20/20 | 20/25 | c.514C>T | p.R172W | Pathogenic | CACD | Mutation at non-covalent oligomerization interface of EC2 | GRM6 c.722-2del, ADGRV1 c.8295G>T (p.L2765F), GNAT2 c.349G>A (p.G117R), MKKS c.1161+4G>A, PRPF4 c.1324G>A (p.V442I), and RP1 c.5522C>T (p.T184I). |
| 3* | 15 - 19 | M | 20/25 | 20/20 | c.514C>T | p.R172W | Pathogenic | CACD | Mutation at non-covalent oligomerization interface of EC2 | GRM6 c.722-2del, MKKS c.1161+4G>A, and REEP6 c.298T>C (p.F100L). |
| 4* | 10 - 14 | F | 20/20 | 20/20 | c.514C>T | p.R172W | Pathogenic | CACD | Mutation at non-covalent oligomerization interface of EC2 | GRM6 c.722-2del, MKKS c.1161+4G>A, PRPF4 c.1324G>A (p.V442I), and REEP6 c.298T>C (p.F100L). |
| 5 | 55 - 59 | F | 20/60 | 20/60 | c.514C>T | p.R172W | Pathogenic | CACD | Mutation at non-covalent oligomerization interface of EC2 | N/A |
| 6 | 25 - 29 | M | 20/20 | 20/25 | c.514C>T | p.R172W | Pathogenic | CACD | Mutation at non-covalent oligomerization interface of EC2 | N/A |
| 7 | 60 - 64 | F | 20/100 | 20/150 | c.514C>T | p.R172W | Pathogenic | CACD | Mutation at non-covalent oligomerization interface of EC2 | N/A |
| 8 | 60 - 64 | M | 20/50 | 20/60 | c.623G>A | p.G208D | Pathogenic | CACD | Mutation at non-covalent oligomerization interface of EC2 | N/A |
| 9 | 35 - 39 | F | 20/20 | 20/30 | c.638G>C | p.C213S | Pathogenic | PD (STG-PD) | Mutation at C166-C213 intramolecular disulfide bridge | N/A |

| ID | Age | Sex | BCVA OD | BCVA OS | Genotype | Proteotype | Classification | Phenotype | Structural outcome of mutations | Other identified variants |
| --- | --- | --- | --- | --- | --- | --- | --- | --- | --- | --- |
| 10 | 75 - 79 | M | CF | 20/30 | c.422A>G | p.Y141C | Pathogenic | CRD | Mutation at covalent oligomerization interface of EC2 | N/A |
| 11 | 55 - 59 | F | 20/50 | 20/125 | c.646C>T | p.P216S | Pathogenic | RP | Mutation at EC2-loop 2 | N/A |
| 12 | 55 - 59 | M | 20/25 | 20/25 | c.403A>G | p.K135E | VUS | CACD | Mutation at non-covalent oligomerization interface of EC2 | N/A |
| 13 | 65 - 69 | M | 20/40 | 20/40 | c.652T>C | p.S218P | VUS | RP | Mutation at EC2-loop 2 | N/A |
| 14# | 75 - 79 | F | 20/100 | 20/125 | c.914G>A | p.G305D | VUS | PD (AFVD) | Mutation at fusion regulatory region of IC3 | N/A |
| 15# | 40 - 44 | F | 20/20 | 20/40 | c.914G>A | p.G305D | VUS | PD (AFVD) | Mutation at fusion regulatory region of IC3 | N/A |
| 16 | 45 - 49 | M | 20/200 | 20/200 | c.623G>A, c.683C>T | p.G208D, p.T228I (unknown whether cis or trans) | Pathogenic, VUS | CACD | Mutation at non-covalent oligomerization interface of EC2 | N/A |
| 17 | 50 - 54 | F | 20/80 | 20/125 | c.548G>A | p.R183H | Novel | RP | Mutation at covalent oligomerization interface of EC2 | N/A |
| 18 | 35 - 39 | M | 20/80 | 20/100 | c.791C>G | p.S264C | Novel | RP | Mutation at TM4 | ABCA4 (c.2666T>C [p.V889A]). |
| **Frameshift and Nonsense Variants** |  |  |  |  |  |  |  |  |  |  |
| 19 | 50 - 54 | M | 20/40 | 20/30 | c.113delG | p.G38Dfs*4 | Pathogenic | CRD | Frameshift begins at TM1 | N/A |
| 20 | 35 - 39 | M | 20/20 | 20/30 | c.136C>T | p.R46* | Pathogenic | CRD | Terminates in EC1 | N/A |
| 21 | 45 - 49 | M | 20/40 | 20/25 | c.303C>G | p.Y101* | Pathogenic | PD (STG-PD) | Terminates in TM3 | N/A |
| 22** | 60 - 64 | M | 20/400 | 20/80 | c.571G>T | p.E191* | Pathogenic | CACD | Terminates in EC2 | N/A |
| 23** | 35 - 39 | M | 20/20 | 20/20 | c.571G>T | p.E191* | Pathogenic | Normal | Terminates in EC2 | N/A |
| 24## | 75 - 79 | F | NA | NA | c.271T>A, c.310delATCT | p.Y91N, p.I104Vfs*34 (likely in cis) | VUS, Pathogenic | PD (STG-PD) | Frameshift begins at TM3 | N/A |
| 25## | 45 - 49 | F | 20/20 | 20/40 | c.271T>A, c.310delATCT | p.Y91N, p.I104Vfs*34 (likely in cis) | VUS, Pathogenic | PD (STG-PD) | Frameshift begins at TM3 | N/A |
| 26## | 30 - 34 | F | 20/20 | 20/20 | c.271T>A, c.310delATCT | p.Y91N, p.I104Vfs*34 (likely in cis) | VUS, Pathogenic | PD (STG-PD) | Frameshift begins at TM3 | N/A |
| 27 | 60 - 64 | F | 20/40 | 20/70 | c.246C>A | p.C82* | VUS | PD (BPD) | Terminates in IC2 | N/A |
| 28 | 70 - 74 | M | 20/40 | 20/40 | c.265dup | p.A89Gfs*88 | Novel | CRD | Frameshift begins at IC2 | N/A |
| 29 | 60 - 64 | F | 20/50 | 20/50 | c.665delC | p.P219Hfs*37 | Novel | CD | Frameshift begins at EC2 | N/A |
| 30 | 60 - 64 | F | 20/40 | 20/25 | c.700_703dup | p.S235Ifs*67 | Novel | PD (AFVD) | Frameshift begins at EC2 | N/A |
| **Insertion Variant** |  |  |  |  |  |  |  |  |  |  |
| 31 | 60 - 64 | M | 20/250 | 20/40 | c.776_777insTCT | p.S259_S260insL | Novel | CRD | In-frame insertion at the end of the EC2 domain | N/A |

Patients with the same symbol (\*, \*\*, #, ##) after the ID are part of the same family. Abbreviations: AFVD (adult-onset foveomacular vitelliform dystrophy), BCVA (best corrected visual acuity), BPD (butterfly pattern dystrophy), CACD (Central Areolar Choroidal Dystrophy), CF (count fingers), OD (right eye), OS (left eye), PD (pattern dystrophy), STG-PD (Stargardt's-like pattern dystrophy), NA (not available).

